# Beyond the 24-Hour Rhythm: Functional Principal Components of Daily Activity Residuals and Mortality in NHANES

**DOI:** 10.64898/2026.11.10.26367785

**Authors:** Xin Zheng

## Abstract

**Background:** Accelerometer-derived 24-hour activity profiles are usually summarised by cosinor rhythm parameters or by functional principal component analysis (FPCA). Applied to the raw profiles, however, the leading functional components largely re-express what the cosinor already captures: activity level, rhythm strength and timing. Whether the shape of the day carries health information beyond these parameters is unclear.

**Methods:** We analysed adults with seven valid days of accelerometry from two National Health and Nutrition Examination Survey (NHANES) cohorts: hip activity counts (2003–2006; *n* = 2,615) and wrist Monitor-Independent Movement Summary units (2011–2014; *n* = 3,208). Minute-level activity was transformed with the inverse hyperbolic sine, and a single 24-hour cosinor curve was fitted across each participant’s seven days. Each participant’s average residual curve was decomposed by FPCA and, to separate stable between-person patterns from day-to-day variation, by multilevel FPCA. The residual components were compared with standard FPCA of the average activity curves. In exploratory analyses, we related the three leading components to all-cause mortality and, secondarily, to cardiovascular and cancer mortality. We used Cox proportional hazards models adjusted for age, sex, ethnicity, income and the cosinor parameters, with the hip cohort for discovery and the wrist cohort for replication.

**Results:** The three leading residual components explained 44% (hip) and 51% (wrist) of residual variance and consisted mainly of 12-hour and 8-hour patterns. The second component (FPC2) contrasts an early-starting, flat day with a day of distinct mid-morning and evening peaks. Its shape was nearly identical in the two cohorts (similarity 0.96), and multilevel FPCA recovered it as a stable between-person pattern. In unweighted models, FPC2 was the only component associated with all-cause mortality in the hip discovery cohort, which replicated in the wrist cohort. With survey weights that also accounted for selection into the analytic sample, the association was concentrated in cardiovascular deaths (hip 1.53, 1.12 to 2.09, *p* = 0.007; wrist 1.38, 1.10 to 1.72, *p* = 0.005). This association persisted after further adjustment for lifestyle factors, traditional activity metrics and self-reported diseases. In standard FPCA, the pattern was mixed with 24-hour rhythm variation, unstable in small samples, and showed no association unless the cosinor parameters were adjusted for. FPC2 was not associated with cancer mortality. Scores of the leading components did not form distinct clusters (average silhouette width *≤* 0.23), but FPC2 was selected as a continuous predictor of cardiovascular mortality in both cohorts by forward selection on 10-fold cross-validated, survey-weighted AUC.

**Conclusions:** The within-day structure of activity that a 24-hour cosinor cannot represent varies continuously between people. One replicated aspect of it, the loss of a two-peak daily pattern, was associated with mortality beyond rhythm strength and timing, most clearly cardiovascular mortality. The shape of daily activity therefore carries prognostic information beyond standard rhythm summaries. It can support mortality risk prediction that needs only a few personal details, and may help identify individuals at elevated risk and tailor interventions to their daily activity patterns.

## 1 Introduction

Accelerometers now record human movement continuously over 24 hours in large population studies, including the National Health and Nutrition Examination Survey (NHANES) and the UK Biobank [1, 2]. Each participant contributes several days of high-resolution data, yet most epidemiological studies reduce these recordings to scalar summaries such as total activity or minutes of moderate-to-vigorous physical activity (MVPA). Functional data analysis (FDA) offers an alternative: by modelling the 24-hour PA profile as a continuous function of time-of-day, it analyses the whole day as a smooth curve [3], accounting for the ordering of the measurements and their temporal correlation [4, 5]. An early example is the functional linear model of Wang et al. [6], who represented averaged activity profiles with a nine-term Fourier basis and related them to sleep apnoea.

Functional principal component analysis (FPCA) has become the most widely used FDA tool for accelerometer data [7, 8]. Applied to 24-hour activity profiles, it identifies a small number of dominant components of variation. In older men in the MrOS study, the first four components predicted changes in sleep, cognition and mortality [9]. In NHANES 2011– 2014, Xiao et al. [10] identified four patterns in U.S. adults, interpreted as high amplitude, early rise, a prolonged activity window and a bi-phasic pattern, and related them to sociodemographic factors and self-rated health; a companion analysis applied the same approach to U.S. adolescents [11]. Because accelerometer data have a nested structure, with days within participants, multilevel FPCA was introduced to separate variation between participants from day-to-day variation within participants [12, 13]. It has since been extended beyond continuous activity profiles to other types of activity profiles, including a generalized multilevel FPCA for binary active/inactive profiles in NHANES [14] and a quantile-based formulation that replaced the mean curve with quantile curves to capture distributional features of activity [15]. In addition, functional clustering groups participants with similar curves into phenotypes [16, 17], an approach that is attractive for describing population heterogeneity and for communicating interpretable behavioral phenotypes to public users of wearable devices. Clustering can also identify combinations of functional characteristics that are difficult to summarize using single components [18, 19].

In parallel, chronobiology summarises the same recordings with a small set of rhythm parameters [20]. The cosinor model [21, 22] describes the day by its rhythm-adjusted mean level (mesor), the strength of the day–night contrast (amplitude) and the timing of the peak (acrophase); these parameters are widely used to characterise the rest–activity rhythm [23, 24, 25]. The extended cosinor adds two parameters for the width and steepness of the active period, but is restricted to shapes that are symmetric around the peak [26]. Besides, nonparametric metrics such as interdaily stability (IS) and intradaily variability (IV) describe further aspects of irregularity in non-sinusoidal PA patterns [27]. Circadian rhythm estimates depend on the method used to compute them [28], and a recent review of these approaches suggested developing a construct that encompasses all characteristics of the rest– activity rhythm [20], without considering FDA.

The two traditions in modeling PA patterns have developed largely separately, yet they overlap more than is usually acknowledged. The 24-hour-periodic Fourier basis used in FDA smoothing decomposes the day into the 24-hour component and its higher harmonics (periods of 12, 8, 6 hours and so on), and the 24-hour component is closely related to the cosinor model. Consistent with this, Xiao et al. [29, 10] showed that the leading components of FPCA on 24-hour profiles largely re-express amplitude and acrophase. In a recent study [30], cosinor and FPCA were run side by side on the same data and compared only by correlation, reporting that the leading FPCs aligned with the cosinor timing metrics. Curve registration can separate timing from the remaining variation [31, 25], but it does so by removing timing, which is itself of health interest. Furthermore, the cosinor assumes a symmetric daily rhythm around its peak [26] which cannot independently represent, for example, different steepness of the morning and evening transitions. As a result, it remains unclear whether the functional representation carries health-relevant information beyond level, 24-hour rhythm strength and timing, or whether it mainly re-expresses these parameters in a less interpretable form.

In this study we address this question directly by combining the two traditions in a decom-position that separates these features by construction. For each participant, a cosinor model captures the typical level, rhythm strength and timing, and the residual curve captures the remaining shape of the day, which is then analysed by FPCA. This yields shape components that do not overlap with the cosinor parameters. Using two NHANES cohorts with different devices, hip-worn activity counts in 2003–2006 and wrist-worn MIMS units in 2011–2014, we (i) describe the main components of shape variation and their overlap with existing rhythm metrics, (ii) explore whether shape components are associated with all-cause mortality, and secondarily with cardiovascular and cancer mortality, beyond level, rhythm strength, timing and activity volume, and (iii) examine whether participants form distinct activity phenotypes by functional clustering. Finally, we provide a web application in which users can upload their own accelerometer data and obtain the same decomposition, their shape score relative to the NHANES cohorts and their nearest activity group.

## 2 Methods

### 2.1 Study population

We used two NHANES cohorts. In 2003–2006, participants wore a hip-worn accelerometer that recorded activity counts per minute; in 2011–2014, participants wore a wrist-worn accelerometer, and minute-level activity was summarised as Monitor-Independent Movement Summary (MIMS) units [32]. Inclusion and exclusion criteria, non-wear handling, and covariate definitions followed Leroux et al. [1] for 2003–2006 and were applied analogously to 2011–2014, as in our previous work [33]. The resulting analytic samples comprised 2,615 participants in the hip-worn cohort (2003–2006) and 3,208 in the wrist-worn cohort (2011– 2014); their characteristics are summarized in Table 1.

**Table 1:** Participant characteristics. Values are mean (SD), median (IQR) or percentage.

|  | Hip counts, 2003–2006 | Wrist MIMS, 2011–2014 |
|---|---|---|
| Participants, *n* | 2,615 | 3,208 |
| Age, years | 55.8 (17.4) | 52.7 (17.4) |
| Female, % | 48.1 | 56.1 |
| ethnicity, % |  |  |
| Non-Hispanic White | 59.4 | 42.6 |
| Non-Hispanic Black | 15.9 | 22.6 |
| Hispanic | 20.3 | 21.3 |
| Other | 4.4 | 13.5 |
| Income-to-poverty ratio < 1.3, % | 21.5 | 33.0 |
| Income-to-poverty ratio ≥ 3.5, % | 37.4 | 32.1 |
| BMI, kg/m<sup>2</sup> | 27.9 (5.5) | 29.3 (7.1) |
| Current smoker, % | 16.6 | 17.2 |
| Current drinker, % | 59.8 | 61.8 |
| More than high school, % | 51.4 | 57.6 |
| Any mobility difficulty, % | 19.9 | 21.7 |
| Mean activity per minute<sup>*a*</sup> | 175.7 (91.6) | 9.1 (2.5) |
| MVPA, min/day<sup>*b*</sup> | 13 (4–28) | 256 (182–337) |
| Wear time, hours/day<sup>*c*</sup> | 15.2 (2.2) | Wake 15.3 (1.3) + Sleep 7.6 (1.4) |
| Cosinor mesor (asinh units) | 2.36 (0.56) | 1.91 (0.27) |
| Cosinor amplitude (asinh units) | 2.24 (0.59) | 1.26 (0.29) |
| Cosinor acrophase, clock time | 14:15 (13:25–15:12) | 14:46 (13:54–15:46) |
| Interdaily stability (IS) | 0.46 (0.13) | 0.59 (0.13) |
| Intradaily variability (IV) | 0.98 (0.32) | 0.72 (0.21) |
| Follow-up, years | 10.6 (9.6–11.7) | 6.8 (5.8–7.8) |
| Deaths, *n* | 463 | 312 |
| Cardiovascular | 112 | 106 |
| Cancer | 101 | 77 |

<sup>*a*</sup>Activity counts per minute (hip) or MIMS units per minute (wrist), not comparable between cohorts.
<sup>*b*</sup>At least 2020 counts per minute (hip) [1] or 19.614 MIMS units per minute (wrist) [34]; the two definitions are not comparable.
<sup>*c*</sup>Hip: total wear time; the device was removed for sleep. Wrist: the device was worn day and night, and wear time is split into wake and sleep wear as classified by NHANES.

For sensitivity analyses (Supplementary Table S1), we also defined an expanded sample that applied only the accelerometry and mortality-linkage criteria, comprising 2,917 and 3,605 participants aged 18 years or older. This sample retains participants with missing BMI, education or laboratory measures, which do not enter the primary models, and avoids excluding survivors by follow-up time, which conditions on the outcome.

### 2.2 Transformation

Activity was transformed with the inverse hyperbolic sine to reduce skewness and peak dominance,

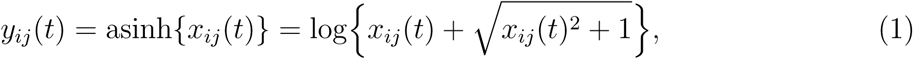

where *x_ij_*(*t*) is the activity of participant *i* = 1*, . . . , n* on day *j* = 1*, . . . ,* 7 at minute *t* of the day. The same transformation was used in both cohorts.

### 2.3 Decomposition of daily activity profiles

For each participant, a 24-hour cosinor model was fitted jointly to all seven days by ordinary least squares, using the R package ActCR [21],

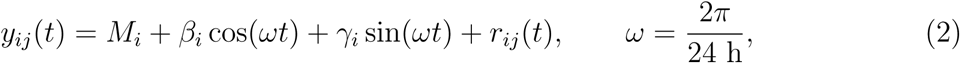

where the mesor *M_i_* is the rhythm-adjusted mean activity level and *r_ij_*(*t*) is the residual. The amplitude and acrophase follow from the two harmonic coefficients,

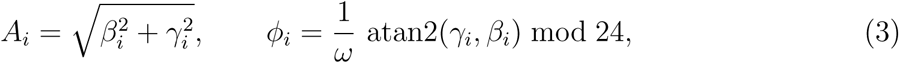

so that the fitted rhythm can be written as *M_i_* + *A_i_* cos*{ω*(*t − ϕ_i_*)*}* The amplitude describes the strength of the day–night contrast, and the acrophase is the clock time of the fitted peak. Day-to-day variability in timing was summarised separately by the circular standard deviation (in hours) of the acrophases obtained from cosinor fits to each day.

For comparison, the extended cosinor model [26],

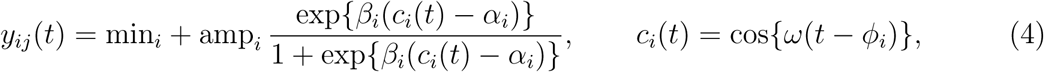

was also fitted to each participant’s seven days with ActExtendCosinor in ActCR, yielding the minimum, amplitude, *α* (relative width of the active period), log *β* (steepness of the rest– activity transitions) and acrophase. These parameters were used only to assess the overlap of shape with existing rhythm metrics and as additional adjustment; the residual shape was derived from the standard cosinor (2).

### 2.4 Functional principal component analysis

The participant’s *shape* is the average residual 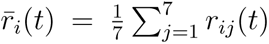. Let *H* = span{1, cos(*ωt*), sin(*ωt*)*}* and let *P_H_* be the orthogonal projection onto *H*. With complete, equally sampled days, the weekly fit equals the cosinor fit to the average day 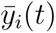, so that

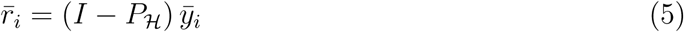

(Supplementary Sections S1.2–S1.3). The shape therefore has zero mean and no 24-hour component, and contains only the 12-hour and higher harmonics of the average day. Level (*M_i_*), rhythm (*A_i_, ϕ_i_*) and shape 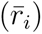 describe non-overlapping parts of each participant’s average day, although they may be correlated across participants. These properties hold exactly because all participants share the same design matrix *X* = [ 1, cos(*ωt*), sin(*ωt*)], defined by a common time grid and a fixed 24-hour period. The projection *P_H_* = *X*(*X^⊤^X*)*^−^*^1^*X^⊤^* is thus identical for all participants, so every residual lies in the same space *H^⊥^*, as do the eigenfunctions estimated from all residuals in the FPCA below (Supplementary Section S1). Supplementary Figure S1 illustrates the decomposition.

#### 2.4.1 Single-level FPCA of participant shape

The participant shapes were decomposed as

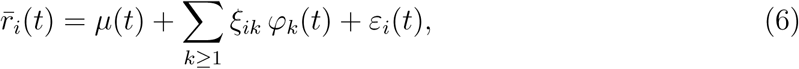

where *µ*(*t*) is the mean shape, *φ_k_* are orthonormal eigenfunctions of the covariance operator 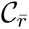 with eigenvalues *λ*_1_ *≥ λ*_2_ *≥ . . .* , *ξ_ik_* are uncorrelated participant scores and *ε_i_*(*t*) is measurement error. By (5), 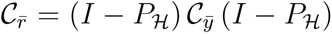, where *C_ȳ_* is the covariance operator of the average days, so every eigenfunction with a positive eigenvalue is orthogonal to the constant and to the 24-hour harmonic. Residual FPCA is therefore not equivalent to FPCA of the activity curves with the leading components discarded, whose eigenfunctions can mix 24-hour and higher-harmonic variation (Supplementary Sections S1.4–S1.6). Orthogonality holds for the eigenfunctions over time, not for the scores across participants: shape scores may still be associated with mesor, amplitude or acrophase (Supplementary Section S1.7), which we therefore included as covariates in the association analyses (Section 2.6.2).

The covariance was estimated with the fast covariance estimation (FACE) method [35], which smooths the covariance with penalised splines (penalty chosen by generalised crossvalidation) and removes the measurement-error contribution, using fpca.face in the R package refund; components were retained up to 90% of explained variance. Component *k* explains a proportion *λ_k_*/∑*_l_λ_l_* of shape variance.

Each component was described by (i) the curves 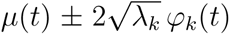; (ii) the observed average day of participants scoring at least 1 SD above and at least 1 SD below the mean; (iii) the share of *φ_k_* at the 24-, 12- and 8-hour periods, as the *R*^2^ from regressing *φ_k_*(*t*) on a cosine and sine of that period; and (iv) the similarity of the hip and wrist eigenfunctions, as the inner product of the normalised curves (1 = identical shape).

#### 2.4.2 Multilevel FPCA

To separate a participant’s usual shape from day-to-day variation, multilevel FPCA [12] was applied to the daily residuals:

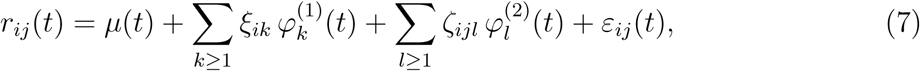

where *φ_k_*^(1)^ and *ξ_ik_* describe between-participant variation (level 1, the usual shape) and *φ_l_*^(2)^ and *ζ_ijl_* describe within-participant variation (level 2, each day’s deviation from it). The model was fitted with mfpca.face in refund [13], retaining components up to 90% of explained variance at each level. Because the cosinor was fitted to the whole week, within-participant deviations equal *y_ij_ − ȳ_i_* and are not projected off *H*; level-2 components can therefore include day-to-day changes in level, amplitude and timing as well as shape, whereas level-1 components describe shape beyond each participant’s own cosinor rhythm (Supplementary Section S1.8). Day-to-day variability was summarised per participant by the SD over the seven days of each of the three leading level-2 scores and by the overall day-to-day variability, (∑*_l_*SD*_i_*(ζ·_l_)2)^1/2^.

#### 2.4.3 Scoring new participants

To score a new participant, we fitted the same weekly cosinor to the participant’s asinh-transformed minute-level activity and averaged the residuals across days to obtain the shape *r̄_i_*(*t*). We then projected this curve, centred at the cohort mean, onto the fitted eigenfunction *φ_k_*. Substituting (6) and using the orthonormality of the eigenfunctions, ∑*_t_φ_k_*(*t*)*φ_l_*(*t*) = **1**(*k* = *l*), gives

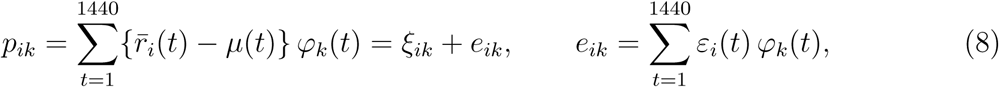

where Var(*ξ_ik_*) = *λ_k_* and *e_ik_* is the measurement error carried into the projection. If *ε_i_*(*t*) is uncorrelated across minutes with variance *σ*^2^, then E(*e_ik_*) = 0 and Var(*e_ik_*) = *σ*^2^. The projection was then multiplied by the FPCA shrinkage factor *c_k_* to give the score,

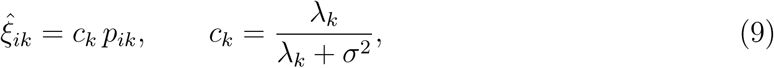

which is the best linear predictor of *ξ_ik_* given *p_ik_*, and equals E(*ξ_ik_ | p_ik_*) when both are normally distributed. The factor *c_k_* is a single constant for each component within a cohort, shared by all participants. The FPC2 score is (9) with *k* = 2. The mean *µ*, eigenfunction *φ_k_*, eigenvalue *λ_k_* and noise variance *σ*^2^ were fixed at their estimates from the fitted cohort; no model was refitted.

#### 2.4.4 Comparison with standard FPCA

To assess what the decomposition adds, we compared the residual components with standard FPCA of the participant-average activity curves *ȳ_i_*(*t*). Each residual component was matched to the standard component with the most similar eigenfunction in the full cohort. The harmonic content of the average days was summarised by their coefficients on a cosine and sine at periods of 24, 12, 8 and 6 hours, and the association between the 24-hour and each shorter-period pair of coefficients across participants by canonical correlation. The stability of the components, which does not involve any outcome, was assessed by re-estimating both FPCAs in participants drawn at random from each cohort, 30 times at each of 300, 500, 1,000 and 2,000 participants, and separately within each of the four two-year NHANES survey waves. For the component carried forward in the mortality analyses, the associations of the residual component and its standard counterpart were compared with and without adjustment for the cosinor parameters.

### 2.5 Functional clustering

To examine whether participants form distinct activity phenotypes, we applied *k*-means clustering to the FPCA scores of the participant-average asinh-transformed activity curves, retaining the components that together explain 90% of variance. Scores were not standardized, so that *k*-means on the scores is equivalent to *L*^2^-based functional clustering of the projected curves. The number of clusters *K* = 1*, . . . ,* 8 was chosen by the gap statistic with the 1-SE rule, which can also indicate the absence of any cluster structure (*K* = 1). Cluster separation was measured by the average silhouette width; values below 0.25 indicate no substantial cluster structure.

### 2.6 Statistical analysis

#### 2.6.1 Rhythm metrics and covariates

Level was represented by the cosinor mesor, rhythm strength by the cosinor amplitude, and timing by the acrophase, entered as the pair *{*cos(*ωϕ_i_*), sin(*ωϕ_i_*)*}*. Shape was represented by the standardised scores of the three leading residual components (FPC1–FPC3), a number fixed before the outcome analyses. Additional rhythm metrics were the extended cosinor minimum, *α* and log *β*; IS and IV from hourly means [27]; the circular SD of the daily acrophases (log-transformed); and the multilevel day-to-day variability summaries. Activity volume was summarised by mean daily wear time, total activity and MVPA. Family income was the income-to-poverty ratio in three categories (*<*1.3, 1.3 to *<*3.5, *≥*3.5), with a fourth category for missing income. Mobility difficulty, BMI, smoking, alcohol use and education followed Leroux et al. [1]. Baseline health status comprised self-reported physician-diagnosed diabetes, congestive heart failure, coronary heart disease, systolic blood pressure, total and HDL cholesterol, and stroke and cancer.

#### 2.6.2 Mortality association analysis

The primary outcome was all-cause mortality; secondary outcomes were cardiovascular and cancer mortality, analysed as cause-specific hazards with deaths from other causes censored. Cox proportional hazards models included age, sex, ethnicity, family income, cosinor mesor, amplitude and acrophase (cosine and sine), and the three shape components jointly (Model 1). Other covariates were included to adjust for potential confounding. Proportional hazards were checked with scaled Schoenfeld residuals. In a sensitivity analysis to reduce the potential influence of reverse causation, the analyses were repeated after excluding participants who died within the first two years of follow-up.

The hip cohort served as the discovery cohort and the wrist cohort as the replication cohort. Testing was hierarchical: all three components were tested only for all-cause mortality in the discovery cohort, with Benjamini–Hochberg correction across FPC1–FPC3, and the component identified there was then tested alone in the replication cohort and for the secondary outcomes. Because this component had the same shape and sign in both cohorts, its cohort-specific log hazard ratios were pooled descriptively by fixed-effect inverse-variance weighting, with heterogeneity assessed by Cochran’s *Q*.

Survey-weighted models were fitted with svycoxph in the R package survey [36], accounting for the sampling weights, strata and primary sampling units, and estimate associations for the US adult population. Unweighted models accounted for the strata and primary sampling units through design-based standard errors but not for the sampling weights, and estimate associations in the study sample conditional on the covariates. Because unweighted estimates are unbiased and more efficient when the weights depend only on covariates in the model [37], both are reported.

#### 2.6.3 Mortality prediction analysis

Given the wide age range in both cohorts and the strong predictive contribution of age, baseline models may already achieve high discrimination [38], leaving limited room for accelerometer-derived features to add discriminative value [39] despite their independent associations with health outcomes [40]. To assess whether the shape component improves prediction of cardiovascular death, we used forward selection among age, sex, ethnicity, income, mesor, amplitude, acrophase and FPC2, following Smirnova et al. [40]. Because NHANES is a complex survey, we used survey-weighted logistic regression rather than machine-learning methods such as random forests or gradient boosting [41], which do not readily accommodate the survey design. Selection was by 10-fold cross-validated, survey-weighted area under the ROC curve (AUC), with survey AIC and EPIC (AIC with a penalty of 4 per parameter) as alternative criteria. In a sensitivity analysis, BMI, smoking, alcohol use, education and mobility difficulty were added as candidate predictors, using the same cross-validation folds and criteria.

#### 2.6.4 Sensitivity analyses

As mentioned above, we conducted four sensitivity analyses: (i) comparing the FPCA components with the level-1 components of a multilevel FPCA, to rule out artifacts of day-to-day variation; (ii) excluding deaths in the first two years of follow-up, to reduce reverse causation; (iii) repeating all analyses in an expanded sample with less restrictive inclusion criteria; and (iv) adding lifestyle and functional variables to the candidate set for forward selection in the mortality prediction models..

## 3 Results

### 3.1 Properties of the decomposition

#### 3.1.1 The cosinor and the structure of its residual

Supplementary Figure S1 shows the decomposition of the daily activity profile for one participant from each cohort. The minute-level residuals were strongly autocorrelated across lags and had heavier tails than a normal distribution (Supplementary Figures S2–S3). The residual therefore contains structured information beyond activity level, rhythm strength and timing.

#### 3.1.2 The leading shape components

After the cosinor was removed, 19 (hip) and 18 (wrist) components were needed to explain 90% of residual variance; the three leading components explained 44.0% and 51.3% (Figure 1, Table 2). None contained any 24-hour content. FPC1 and FPC2 consisted mainly of 12-hour patterns (65% to 86% of their shape), the two-peak structure that a single 24-hour cosine cannot represent, and FPC3 mainly of 8-hour patterns (66% to 72%). FPC1 and FPC2 had nearly identical shapes in the two cohorts (similarity 0.95 and 0.96), whereas FPC3 was only partly similar (0.68).

**Figure 1:**
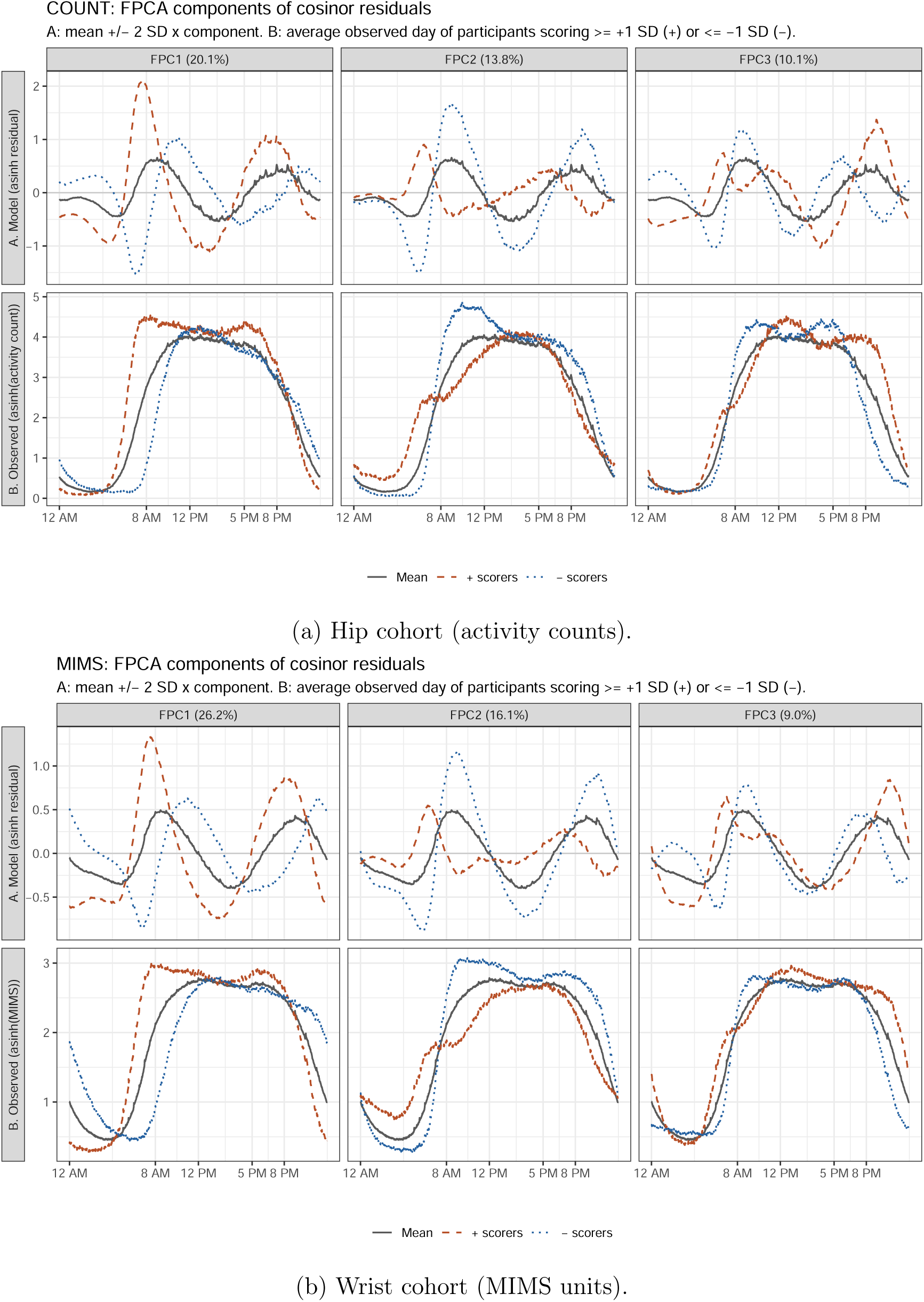
Leading shape components. (A) Model-based mean shape 2 SD along each component. (B) Observed average day of participants scoring at least 1 SD above (red, dashed) or below (blue, dotted) the mean, and of all participants (grey).

**Table 2:**
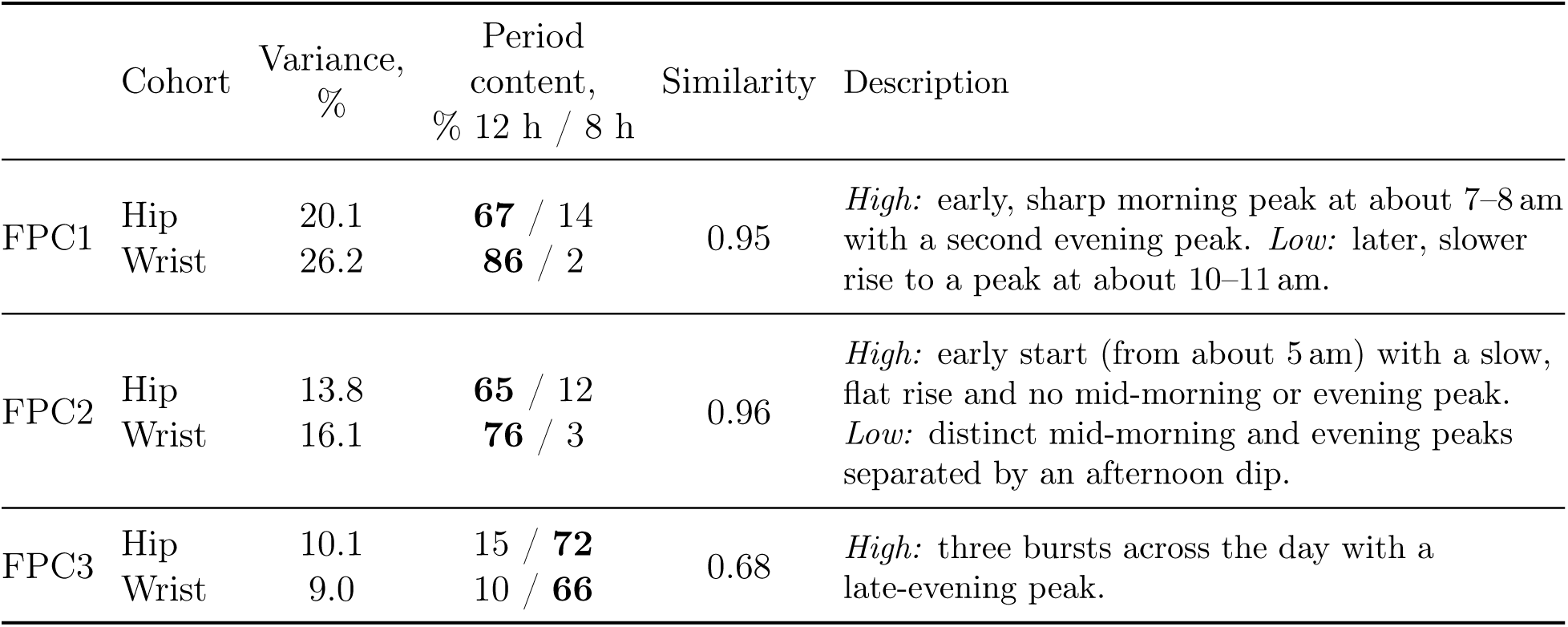
The three leading components of the average residual curve. Variance: share of residual variance. Period content: share of each eigenfunction explained by a 12- or 8-hour cosine and sine. Similarity: inner product of the normalised hip and wrist eigenfunctions.

|  | Cohort | Variance,<br>% | Period<br>content,<br>% 12 h / 8 h | Similarity | Description |
|---|---|---|---|---|---|
| FPC1 | Hip | 20.1 | **67** / 14 | 0.95 | *High:* early, sharp morning peak at about 7–8 am with a second evening peak. *Low:* later, slower rise to a peak at about 10–11 am. |
|  | Wrist | 26.2 | **86** / 2 |  |  |
| FPC2 | Hip | 13.8 | **65** / 12 | 0.96 | *High:* early start (from about 5 am) with a slow, flat rise and no mid-morning or evening peak. *Low:* distinct mid-morning and evening peaks separated by an afternoon dip. |
|  | Wrist | 16.1 | **76** / 3 |  |  |
| FPC3 | Hip | 10.1 | 15 / **72** | 0.68 | *High:* three bursts across the day with a late-evening peak. |
|  | Wrist | 9.0 | 10 / **66** |  |  |

Figures 1a and 1b show the components. FPC1 describes when the morning peak occurs: a sharp peak at about 7–8 am followed by an evening peak, against a later peak at about 10– 11 am. FPC2 describes whether the day has two peaks: high scores correspond to an early, slow start and a flat day, low scores to distinct mid-morning and evening peaks separated by an afternoon dip. FPC3 describes three bursts of activity across the day. As the correlation plot shows (Figure 4), the components were mostly not correlated with existing metrics. The exception was FPC1, which correlated with the cosinor acrophase (Spearman *ρ* = *−*0.38 in the hip cohort and *−*0.65 in the wrist cohort) and with the extended cosinor width *α* (*ρ* = *−*0.45 and *−*0.24). FPC2 correlated weakly with all of them. The absolute correlations were at most 0.22 with the cosinor parameters, IS, IV, total activity and MVPA, and at most 0.27 with the extended cosinor parameters. FPC2 therefore captures a pattern that the existing metrics do not.

In the sensitivity analysis using multilevel FPCA (Figures 2, Supplementary Figures S4 to Supplementary Figures S6) showed that the between-participant (level-1) part accounted for 12.0% (hip) and 14.9% (wrist) of residual variance. The day-to-day (level-2) variation was spread over many components (22 hip, 26 wrist), and the three leading ones explained only 36.4% and 31.0% of it, so no single dominant day-to-day pattern emerged. The leading level-1 eigenfunctions nevertheless recovered the same components as the single-level FPCA (similarity 0.98 to 1.00), and the level-1 and single-level FPC2 scores were highly correlated (0.91 and 0.94), suggesting that the FPC1–FPC3 patterns reflect stable between-participant differences rather than artefacts of day-to-day variation.

**Figure 2:**
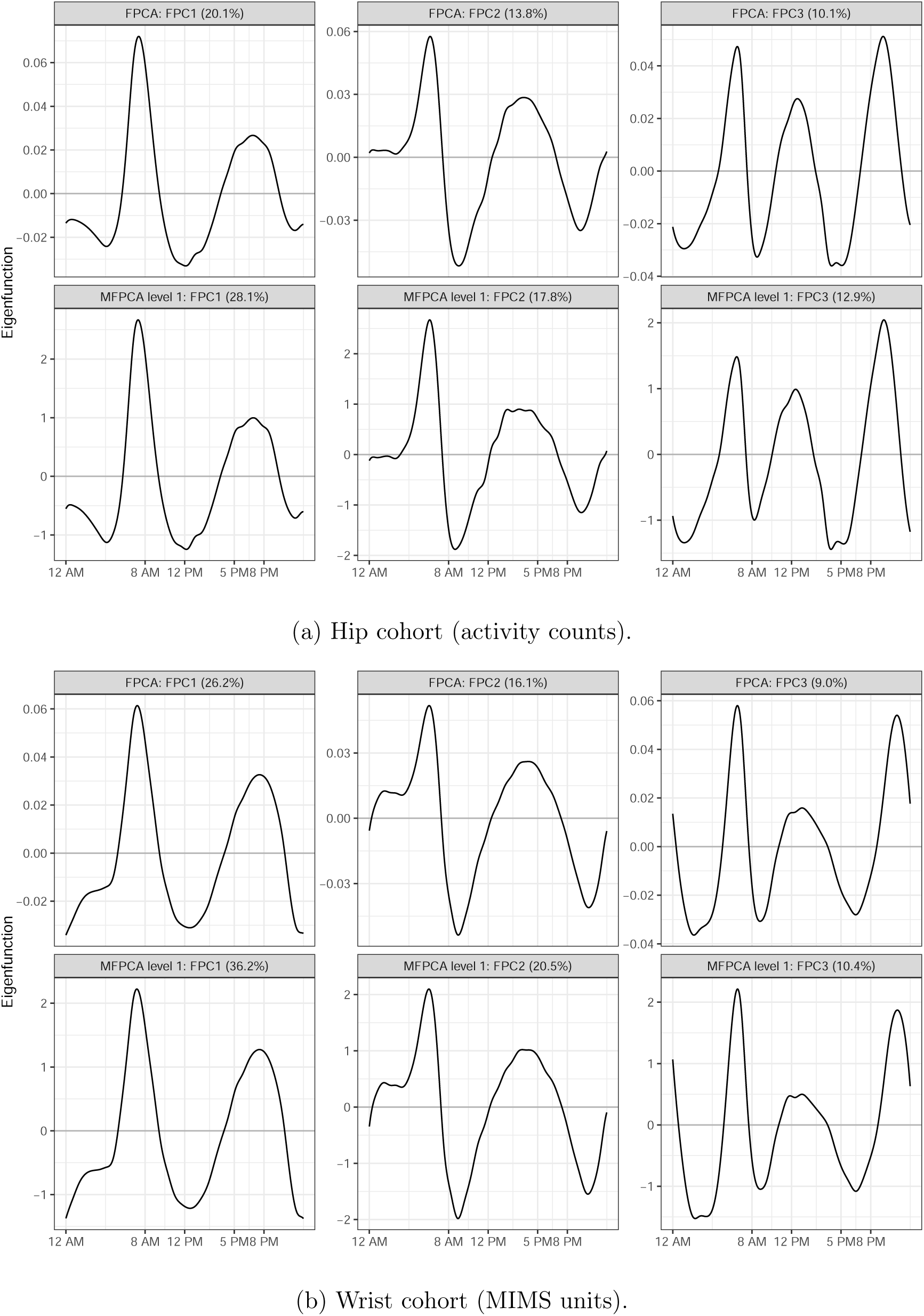
Eigenfunctions of the three Leading shape components.

#### 3.1.3 Comparison with standard FPCA

In Fourier terms, the cosinor fit is the 24-hour harmonic of the average day, and the residual contains the 12-hour and shorter harmonics. Three features of these harmonics explain how standard FPCA of the average curves differs (Table 3A). First, the 24-hour harmonic dominated between-participant variance, so standard FPC1 to FPC3 were mainly level and 24-hour patterns. Second, the cosine and sine directions of the 12-hour harmonic had similar variance, so standard FPC4 and FPC5 were nearly tied (4.4% to 5.1% of variance each). Third, the 24-hour coefficients were correlated with those of the shorter harmonics across participants (canonical correlation 0.25 to 0.68), so standard components mixed 24-hour and shape variation. Removing the 24-hour harmonic before FPCA removes this coupling.

**Table 3:**
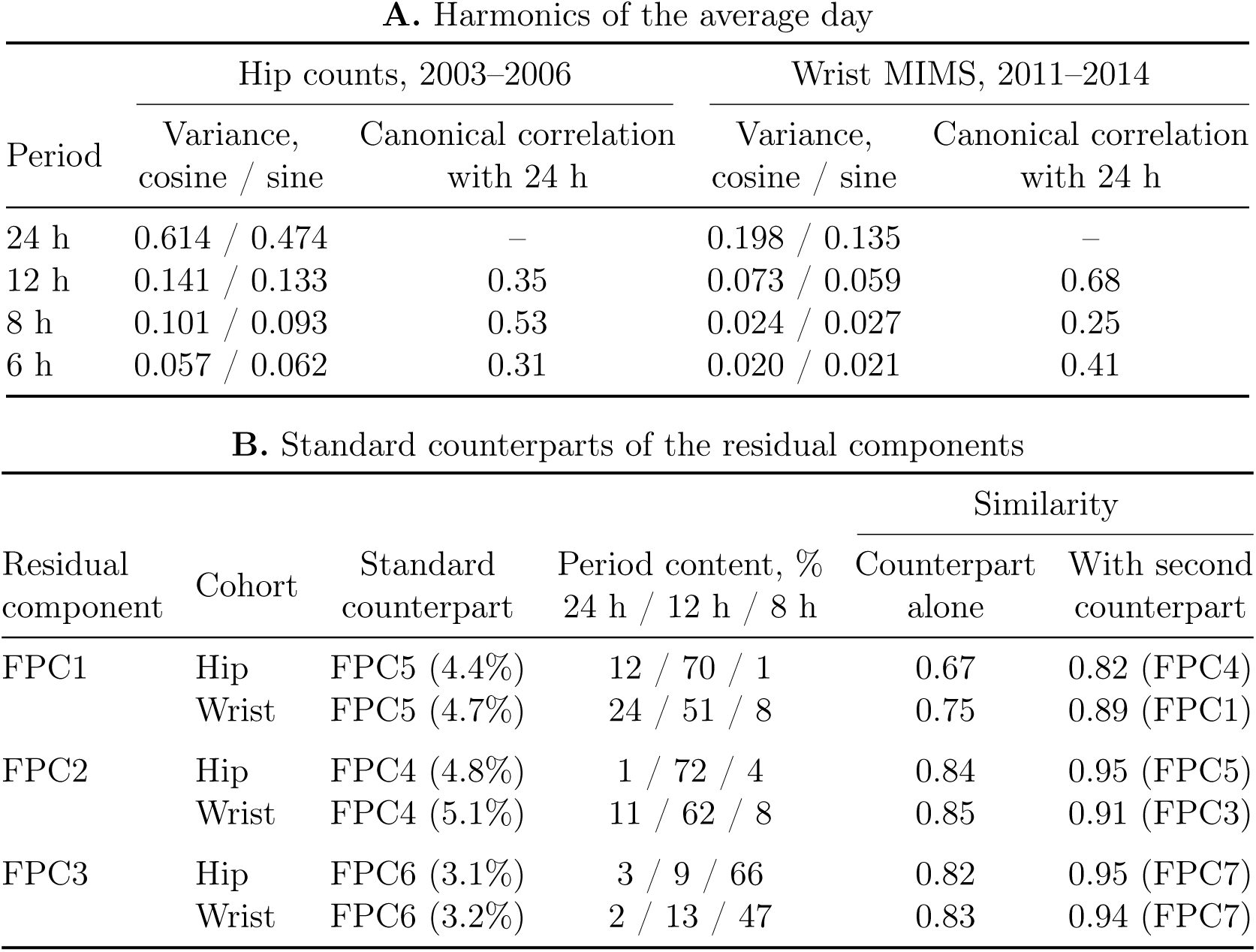
Harmonic structure of the average day and correspondence between residual and standard components. (A) Between-participant variance of the cosine and sine coefficients of each harmonic, and their canonical correlation with the 24-hour coefficients. (B) Standard component with the most similar eigenfunction to each residual component, its period content (share of the eigenfunction explained by a cosine and sine at 24, 12 and 8 hours; the residual components contain no 24-hour content), and the similarity with this component alone and with the subspace of the two most similar standard components.

| **A.** Harmonics of the average day |  |  |  |  |  |
|---|---|---|---|---|---|
|  | Hip counts, 2003–2006 |  | Wrist MIMS, 2011–2014 |  |  |
| Period | Variance, cosine / sine | Canonical correlation with 24 h | Variance, cosine / sine | Canonical correlation with 24 h |  |
| 24 h | 0.614 / 0.474 | – | 0.198 / 0.135 | – |  |
| 12 h | 0.141 / 0.133 | 0.35 | 0.073 / 0.059 | 0.68 |  |
| 8 h | 0.101 / 0.093 | 0.53 | 0.024 / 0.027 | 0.25 |  |
| 6 h | 0.057 / 0.062 | 0.31 | 0.020 / 0.021 | 0.41 |  |
| **B.** Standard counterparts of the residual components |  |  |  |  |  |
|  |  |  |  | Similarity |  |
| Residual component | Cohort | Standard counterpart | Period content, %<br>24 h / 12 h / 8 h | Counterpart alone | With second counterpart |
| FPC1 | Hip | FPC5 (4.4%) | 12 / 70 / 1 | 0.67 | 0.82 (FPC4) |
|  | Wrist | FPC5 (4.7%) | 24 / 51 / 8 | 0.75 | 0.89 (FPC1) |
| FPC2 | Hip | FPC4 (4.8%) | 1 / 72 / 4 | 0.84 | 0.95 (FPC5) |
|  | Wrist | FPC4 (5.1%) | 11 / 62 / 8 | 0.85 | 0.91 (FPC3) |
| FPC3 | Hip | FPC6 (3.1%) | 3 / 9 / 66 | 0.82 | 0.95 (FPC7) |
|  | Wrist | FPC6 (3.2%) | 2 / 13 / 47 | 0.83 | 0.94 (FPC7) |

As a result, each residual component appeared in standard FPCA only at a lower rank, only partly (similarity 0.67 to 0.85) and with added 24-hour content (Table 3B). In random samples of 300 participants, the residual components were reproduced with median similarity 0.95 to 0.99, whereas the standard components were markedly unstable (Figure 3; Supplementary Figure S11). Residual FPCA thus recovers the shape components reliably even in small samples, the setting of many accelerometry studies. With larger samples, the instability of the standard components decreased. At the size of the NHANES cohorts, the two methods therefore differ mainly in what their components contain (Section 3.2.4).

**Figure 3:**
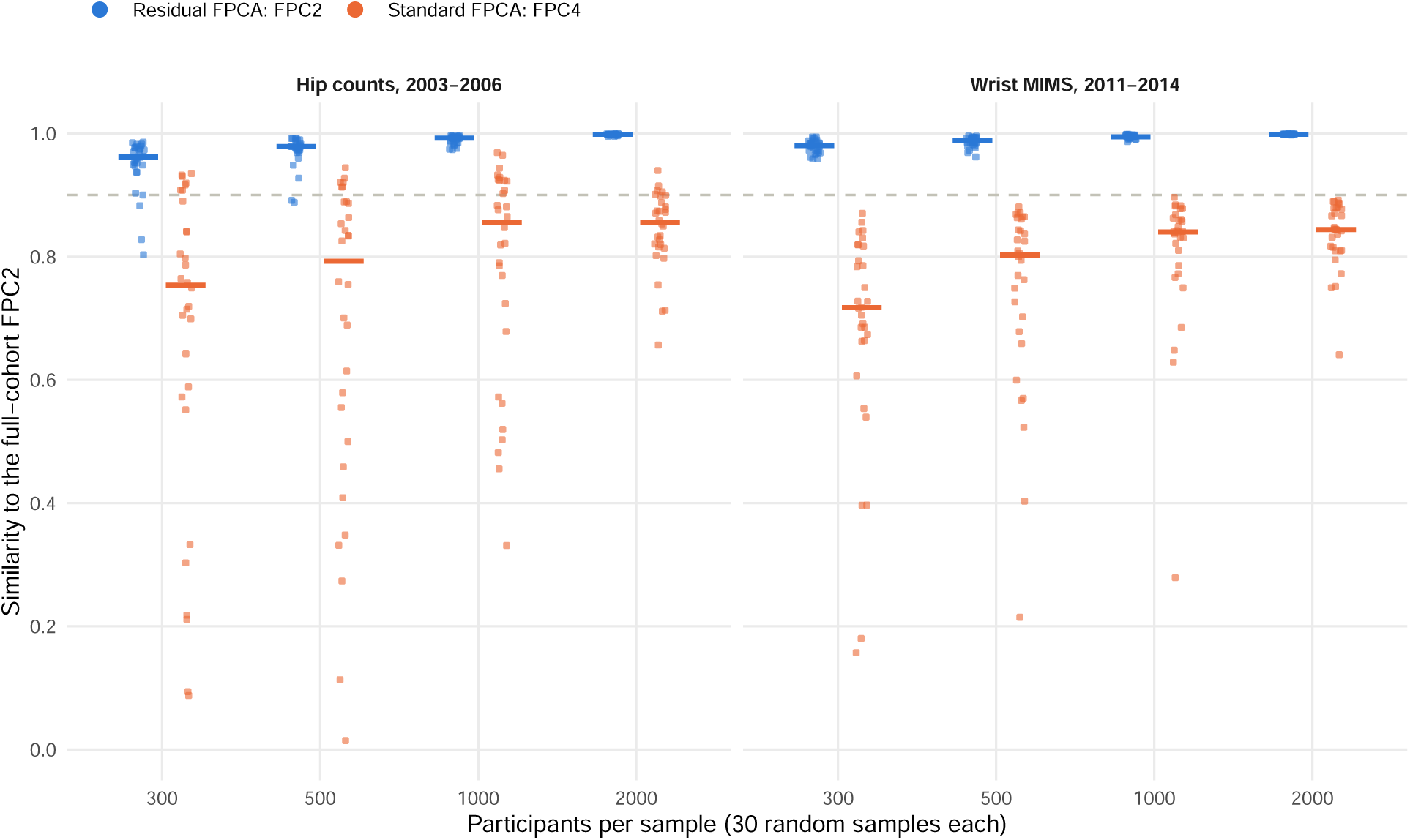
Stability of residual FPC2 and its standard counterpart (standard FPC4) in random samples (FPC1 and FPC3: Supplementary Figure S11). Points: individual samples; bars: median; dashed line: similarity 0.9.

### 3.2 Shape components and mortality

#### 3.2.1 Associations with mortality

In the hip discovery cohort, FPC2 was the only component associated with all-cause mortality (Table 4): in unweighted models with design-based standard errors, the hazard ratio per SD was 1.17 (95% CI 1.06 to 1.29, Benjamini–Hochberg *q* = 0.008), against 1.08 (0.93 to 1.25) for FPC1 and 1.04 (0.92 to 1.18) for FPC3. FPC2 was therefore carried forward and was replicated in the wrist cohort (1.18, 1.03 to 1.35, *p* = 0.017). There was no evidence against proportional hazards for FPC2 (*p ≥* 0.63).

**Table 4:** Model 1 hazard ratios per SD of the shape components, unweighted, with design-based standard errors, main analytic sample.

|  | Hip counts, 2003–2006 |  |  | Wrist MIMS, 2011–2014 |  |  | Pooled |  |  |
|---|---|---|---|---|---|---|---|---|---|
|  | HR (95% CI) | *p* | PH *p* | HR (95% CI) | *p* | PH *p* | HR (95% CI) | *p* | Het. *p* |
| *All-cause mortality* |  |  |  |  |  |  |  |  |  |
| Deaths / *n* | 463 / 2,615 |  |  | 312 / 3,208 |  |  | 775 / 5,823 |  |  |
| FPC1 | 1.08 (0.93–1.25) | 0.321 (*q*=0.482) | 0.722 | 0.92 (0.78–1.07) | 0.283 | 0.369 |  |  |  |
| FPC2 | **1.17 (1.06–1.29)** | **0.003 (*q*=0.008)** | 0.632 | **1.18 (1.03–1.35)** | **0.016** | 0.671 | **1.17 (1.08–1.27)** | **<0.001** | 0.902 |
| FPC3 | 1.04 (0.92–1.18) | 0.493 (*q*=0.493) | 0.090 | 0.96 (0.83–1.10) | 0.558 | 0.022 |  |  |  |
| *Cardiovascular mortality* |  |  |  |  |  |  |  |  |  |
| Deaths / *n* | 112 / 2,615 |  |  | 106 / 3,208 |  |  | 218 / 5,823 |  |  |
| FPC1 | 1.11 (0.89–1.39) | 0.369 | 0.231 | 0.94 (0.71–1.24) | 0.658 | 0.848 |  |  |  |
| FPC2 | 1.18 (0.89–1.57) | 0.259 | 0.736 | **1.28 (1.06–1.54)** | **0.011** | 0.733 | **1.25 (1.07–1.46)** | **0.006** | 0.647 |
| FPC3 | 0.87 (0.66–1.15) | 0.331 | 0.593 | 0.91 (0.72–1.14) | 0.395 | 0.093 |  |  |  |
| *Cancer mortality* |  |  |  |  |  |  |  |  |  |
| Deaths / *n* | 101 / 2,615 |  |  | 77 / 3,208 |  |  | 178 / 5,823 |  |  |
| FPC1 | 1.08 (0.82–1.43) | 0.566 | 0.647 | 1.25 (0.88–1.76) | 0.210 | 0.208 |  |  |  |
| FPC2 | 0.96 (0.80–1.14) | 0.637 | 0.190 | 1.11 (0.80–1.54) | 0.532 | 0.873 | 0.99 (0.85–1.16) | 0.905 | 0.439 |
| FPC3 | 1.11 (0.83–1.49) | 0.475 | 0.580 | 1.10 (0.81–1.51) | 0.540 | 0.201 |  |  |  |

Models adjusted for age, sex, ethnicity, income, cosinor mesor, amplitude and acrophase.
*q*: Benjamini–Hochberg adjusted over FPC1–FPC3 in the hip discovery cohort for all-cause mortality.
Pooled: fixed-effect inverse-variance estimate for FPC2;
PH *p*: Schoenfeld residual test. Het. *p*: Cochran's *Q* test.

**Table 5:** Model 1 hazard ratios per SD of the shape components, survey-weighted, main analytic sample.

|  | Hip counts, 2003–2006 |  |  | Wrist MIMS, 2011–2014 |  |  | Pooled |  |  |
|---|---|---|---|---|---|---|---|---|---|
|  | HR (95% CI) | *p* | PH *p* | HR (95% CI) | *p* | PH *p* | HR (95% CI) | *p* | Het. *p* |
| *All-cause mortality* |  |  |  |  |  |  |  |  |  |
| Deaths / *n* | 463 / 2,615 |  |  | 312 / 3,208 |  |  | 775 / 5,823 |  |  |
| FPC1 | 1.07 (0.89–1.29) | 0.468 (*q*=0.468) | 0.722 | 0.86 (0.69–1.08) | 0.198 | 0.369 |  |  |  |
| FPC2 | 1.14 (0.99–1.31) | 0.068 (*q*=0.203) | 0.632 | 1.14 (0.93–1.38) | 0.202 | 0.671 | **1.14 (1.02–1.28)** | **0.026** | 0.983 |
| FPC3 | 0.92 (0.80–1.07) | 0.289 (*q*=0.434) | 0.090 | 0.94 (0.79–1.11) | 0.479 | 0.022 |  |  |  |
| *Cardiovascular mortality* |  |  |  |  |  |  |  |  |  |
| Deaths / *n* | 112 / 2,615 |  |  | 106 / 3,208 |  |  | 218 / 5,823 |  |  |
| FPC1 | 1.20 (0.91–1.56) | 0.195 | 0.231 | 0.83 (0.58–1.19) | 0.314 | 0.848 |  |  |  |
| FPC2 | **1.53 (1.12–2.09)** | **0.007** | 0.736 | **1.38 (1.10–1.72)** | **0.005** | 0.733 | **1.43 (1.19–1.71)** | **<0.001** | 0.593 |
| FPC3 | **0.65 (0.48–0.87)** | **0.005** | 0.593 | 0.98 (0.76–1.27) | 0.881 | 0.093 |  |  |  |
| *Cancer mortality* |  |  |  |  |  |  |  |  |  |
| Deaths / *n* | 101 / 2,615 |  |  | 77 / 3,208 |  |  | 178 / 5,823 |  |  |
| FPC1 | 1.24 (0.82–1.86) | 0.310 | 0.647 | 0.98 (0.67–1.44) | 0.922 | 0.208 |  |  |  |
| FPC2 | 0.87 (0.69–1.09) | 0.221 | 0.190 | 1.18 (0.85–1.64) | 0.327 | 0.873 | 0.96 (0.79–1.16) | 0.655 | 0.133 |
| FPC3 | 1.21 (0.78–1.85) | 0.394 | 0.580 | 0.99 (0.63–1.56) | 0.977 | 0.201 |  |  |  |

Models adjusted for age, sex, ethnicity, income, cosinor mesor, amplitude and acrophase.
*q*: Benjamini–Hochberg adjusted over FPC1–FPC3 in the hip discovery cohort for all-cause mortality.
Pooled: fixed-effect inverse-variance estimate for FPC2;
PH *p*: Schoenfeld residual test. Het. *p*: Cochran's *Q* test.

In survey-weighted models, higher FPC2 scores were associated with higher all-cause mortality in both cohorts, with similar but not statistically significant estimates: 1.14 (0.99 to 1.31, *p* = 0.068) in the hip cohort and 1.14 (0.93 to 1.38, *p* = 0.202) in the wrist cohort. Pooled across the two cohorts, the survey-weighted hazard ratio was 1.14 (1.02 to 1.28, *p* = 0.026; heterogeneity *p* = 0.98). The association was concentrated in cardiovascular deaths in survey-weighted models: each SD higher FPC2 score was associated with a hazard ratio for cardiovascular mortality of 1.53 (1.12 to 2.09, *p* = 0.007) in the hip cohort and 1.38 (1.10 to 1.72, *p* = 0.005) in the wrist cohort (pooled 1.43, 1.19 to 1.71). In unweighted models, FPC2 was associated with cardiovascular mortality in the wrist cohort but not in the hip cohort. FPC2 was not associated with cancer mortality. FPC1 was not associated with any outcome.

#### 3.2.2 Interpretation of FPC2

Table 6 compares participants with low, middle and high FPC2 scores, and the FPC2 columns of Figures 1a and 1b show their observed average days. Participants with high scores, the higher-risk end, were more active during night, started their day earlier (from about 4– 6 am), lacked the mid-morning surge seen in the rest of the population and were less active in the evening. Their median cosinor acrophase was 40 (hip) and 63 (wrist) minutes earlier than that of low scorers, and their IS significantly lower. Participants with low scores rose sharply at about 7 am to a strong mid-morning peak at 9–10 am and remained active into the evening.

**Table 6:** Characteristics by FPC2 score: low (≤ -1 SD), middle and high (≥ +1 SD). Values are means, medians (MVPA, acrophase) or percentages. Died: during follow-up.

|  | Hip |  |  |  |  | Wrist |  |  |  |  |
|---|---|---|---|---|---|---|---|---|---|---|
|  | Low | Middle | High | *p*<sup>*c*</sup> | *p*<sup>*d*</sup><sub>age</sub> | Low | Middle | High | *p*<sup>*c*</sup> | *p*<sup>*d*</sup><sub>age</sub> |
| *n* | 403 | 1,802 | 410 |  |  | 491 | 2,197 | 520 |  |  |
| Age, years | 55.8 | 56.6 | 52.1 | **0.002** | – | 51.8 | 53.0 | 52.2 | 0.710 | – |
| Female, % | 54.3 | 49.0 | 38.0 | <**0.001** | <**0.001** | 69.2 | 54.3 | 51.3 | <**0.001** | <**0.001** |
| Income-to-poverty, % |  |  |  |  |  |  |  |  |  |  |
| < 1.3 | 22.0 | 21.8 | 20.2 | 0.545 | 0.591 | 31.8 | 31.9 | 38.2 | **0.041** | **0.040** |
| ≥ 3.5 | 35.7 | 38.0 | 36.7 | 0.771 | 0.947 | 33.0 | 33.5 | 25.2 | **0.009** | **0.010** |
| Mobility difficulty, % | 15.4 | 21.3 | 18.3 | 0.268 | **0.012** | 15.7 | 22.7 | 23.5 | **0.002** | <**0.001** |
| Activity per minute<sup>*a*</sup> | 208 | 165 | 189 | **0.003** | <**0.001** | 10.2 | 8.9 | 8.8 | <**0.001** | <**0.001** |
| MVPA, min/day<sup>*b*</sup> | 19 | 12 | 16 | **0.037** | **0.001** | 312 | 246 | 236 | <**0.001** | <**0.001** |
| Cosinor mesor | 2.64 | 2.28 | 2.42 | <**0.001** | <**0.001** | 2.05 | 1.88 | 1.87 | <**0.001** | <**0.001** |
| Cosinor amplitude | 2.41 | 2.21 | 2.19 | <**0.001** | <**0.001** | 1.35 | 1.26 | 1.19 | <**0.001** | <**0.001** |
| Cosinor acrophase | 14:19 | 14:18 | 13:39 | <**0.001** | <**0.001** | 15:02 | 14:41 | 13:59 | <**0.001** | <**0.001** |
| Interdaily stability | 0.48 | 0.46 | 0.44 | <**0.001** | <**0.001** | 0.64 | 0.58 | 0.56 | <**0.001** | <**0.001** |
| Intradaily variability | 0.98 | 0.99 | 0.94 | **0.037** | 0.083 | 0.70 | 0.72 | 0.73 | **0.012** | **0.014** |
| Died, % | 11.4 | 19.3 | 17.1 | **0.021** | <**0.001** | 5.7 | 10.5 | 10.4 | **0.006** | **0.002** |

<sup>*a*</sup>Counts per minute (hip) or MIMS units per minute (wrist).
<sup>*b*</sup>Definitions differ between cohorts (Table 1).
<sup>*c*</sup>High versus low, unadjusted. <sup>*d*</sup>High versus low, adjusted for age.

High scorers were less active overall than low scorers (189 versus 208 counts per minute, *p* = 0.003; 8.8 versus 10.2 MIMS units per minute, *p <* 0.001), although FPC2 was only weakly correlated with total activity (*ρ* = *−*0.11 and *−*0.21) and MVPA (*−*0.07 and *−*0.19). In both cohorts, high scorers were more often male (38.0% versus 54.3% female in the hip cohort; 51.3% versus 69.2% in the wrist cohort). In the hip cohort, high scorers were younger than low scorers and did not differ in income. In the wrist cohort, the two groups did not differ in age, but high scorers more often had low income. The pattern is therefore not explained by older age or lower income alone. Both are adjusted for in the mortality models for FPC2 (Section 3.2.3).

#### 3.2.3 Robustness of the FPC2–mortality association

Spearman correlations (Figure 4) among the variables of the mortality models show that total activity, MVPA and wear time were highly correlated with each other (*ρ* 0.88 to 0.97) and with the cosinor mesor (*ρ* up to 0.93); they were therefore added to Model 2 one at a time and not to Model 1. Supplementary Figure S7 adds the correlations among the multilevel day-to-day variability summaries.

**Figure 4:**
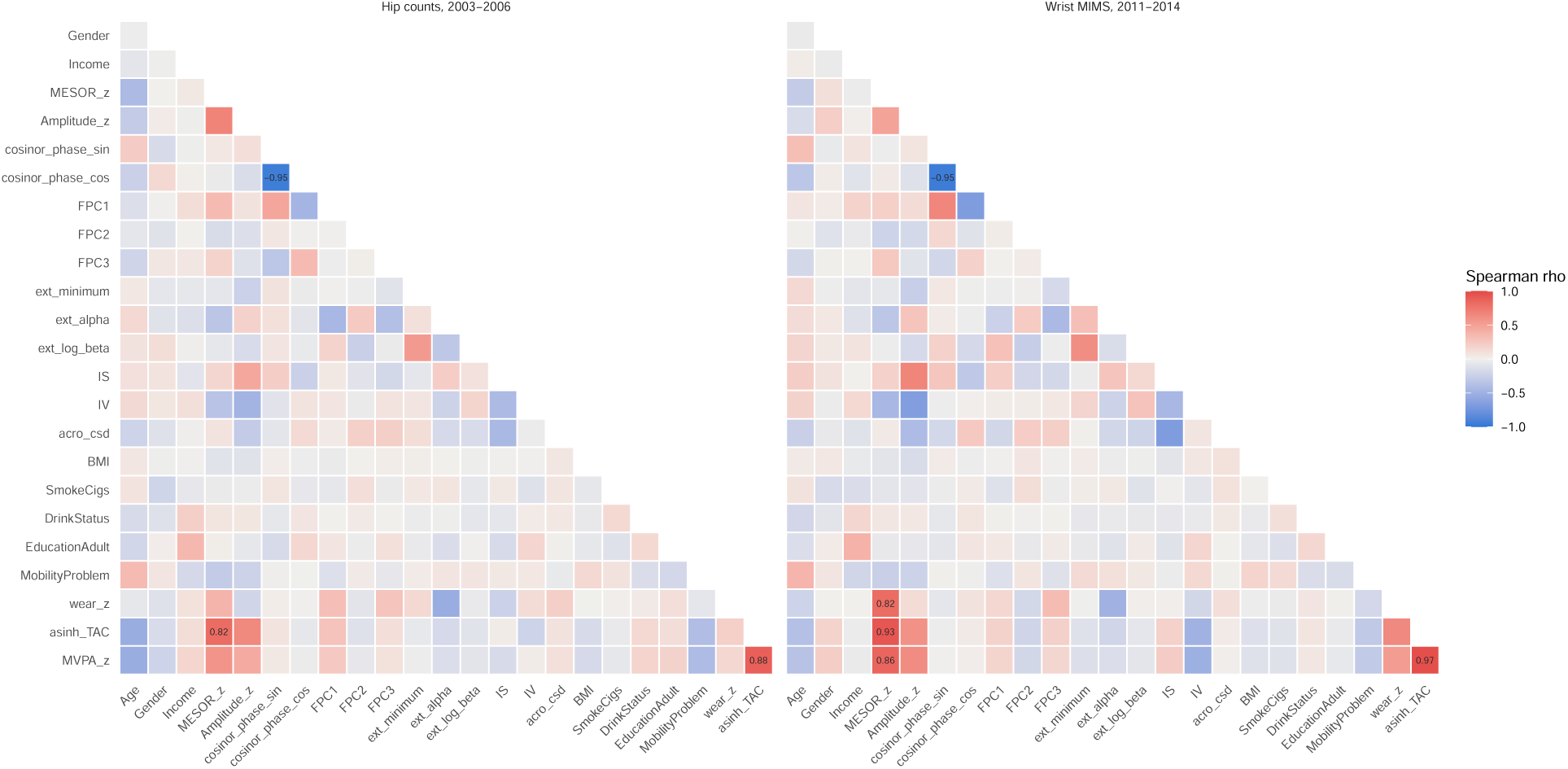
Spearman correlations among the Model 1 and Model 2 covariates, lifestyle factors and activity volume. Coefficients with *|ρ| ≥* 0.7 are printed.

In unweighted models with design-based standard errors (Figure 5), FPC2 remained associated with all-cause mortality in the hip discovery cohort (1.14 to 1.20), and the association was slightly stronger in the wrist replication cohort (1.17 to 1.25). In survey-weighted models, the association of FPC2 with cardiovascular mortality was stable across adjustment sets (Figure 6). Across the 14 specifications, hazard ratios ranged from 1.34 to 1.55 in the hip cohort and from 1.38 to 1.52 in the wrist cohort, and all were statistically significant. In Model 2, which added the extended cosinor parameters, IS, IV, the circular SD of the acrophase and mobility difficulty, the hazard ratios were 1.43 (1.16 to 1.76) and 1.48 (1.13 to 1.94). Further adjustment for wear time, MVPA, BMI, smoking, alcohol and education together gave 1.34 (1.03 to 1.75) and 1.50 (1.16 to 1.93). Adjustment for baseline health status (self-reported congestive heart failure, coronary heart disease, stroke, diabetes and cancer, systolic blood pressure, and total and HDL cholesterol) also left the association unchanged: 1.53 (1.21 to 1.94) and 1.42 (1.13 to 1.79) when added to Model 1, and 1.45 (1.16 to 1.82) and 1.45 (1.11 to 1.90) when added to Model 2.

**Figure 5:**
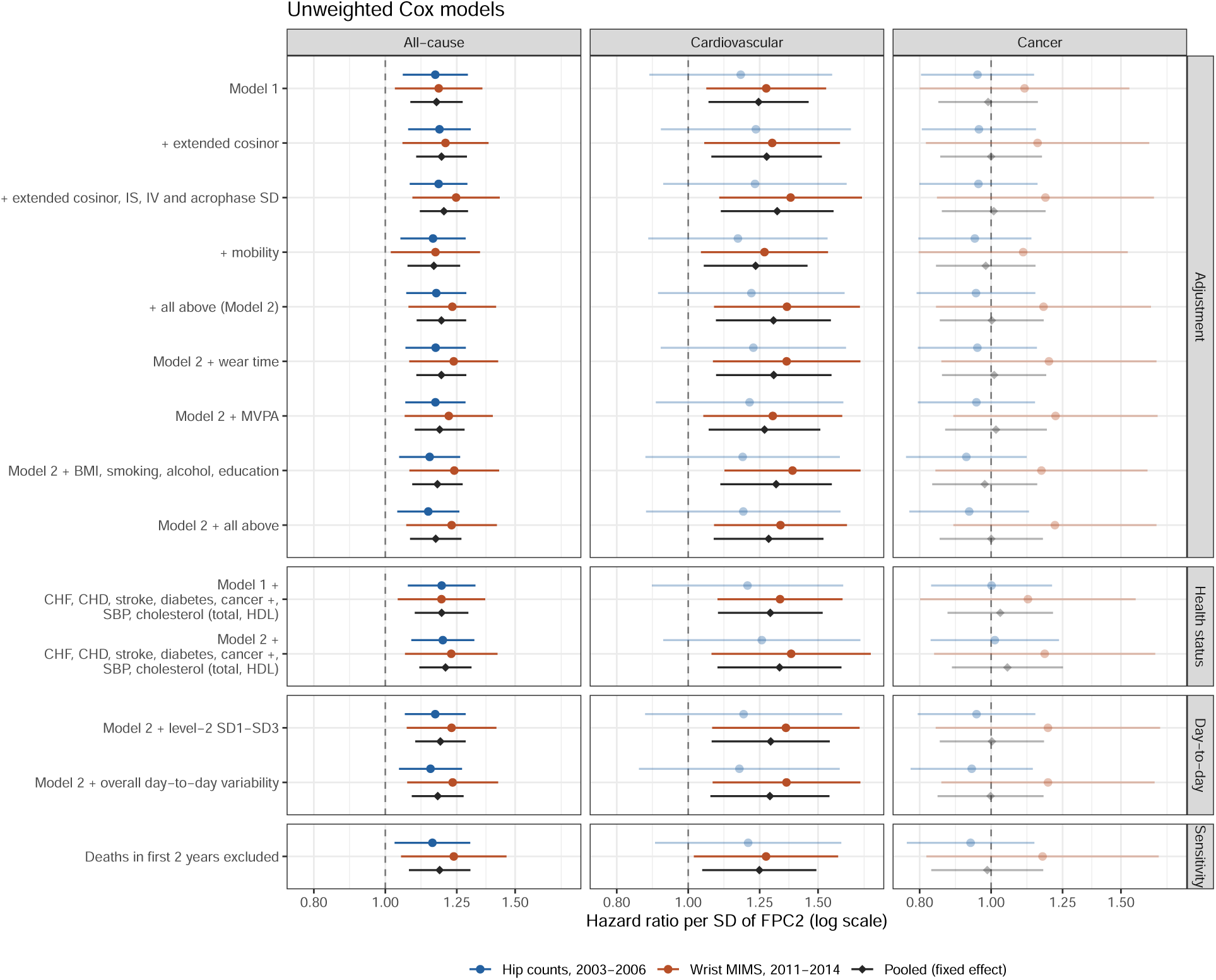
Unweighted hazard ratios for all-cause, cardiovascular and cancer mortality per SD of FPC2 under additional adjustments. Model 1: age, sex, ethnicity, income, cosinor mesor, amplitude and acrophase, and FPC1–FPC3. Faded estimates: *p ≥* 0.05.

**Figure 6:**
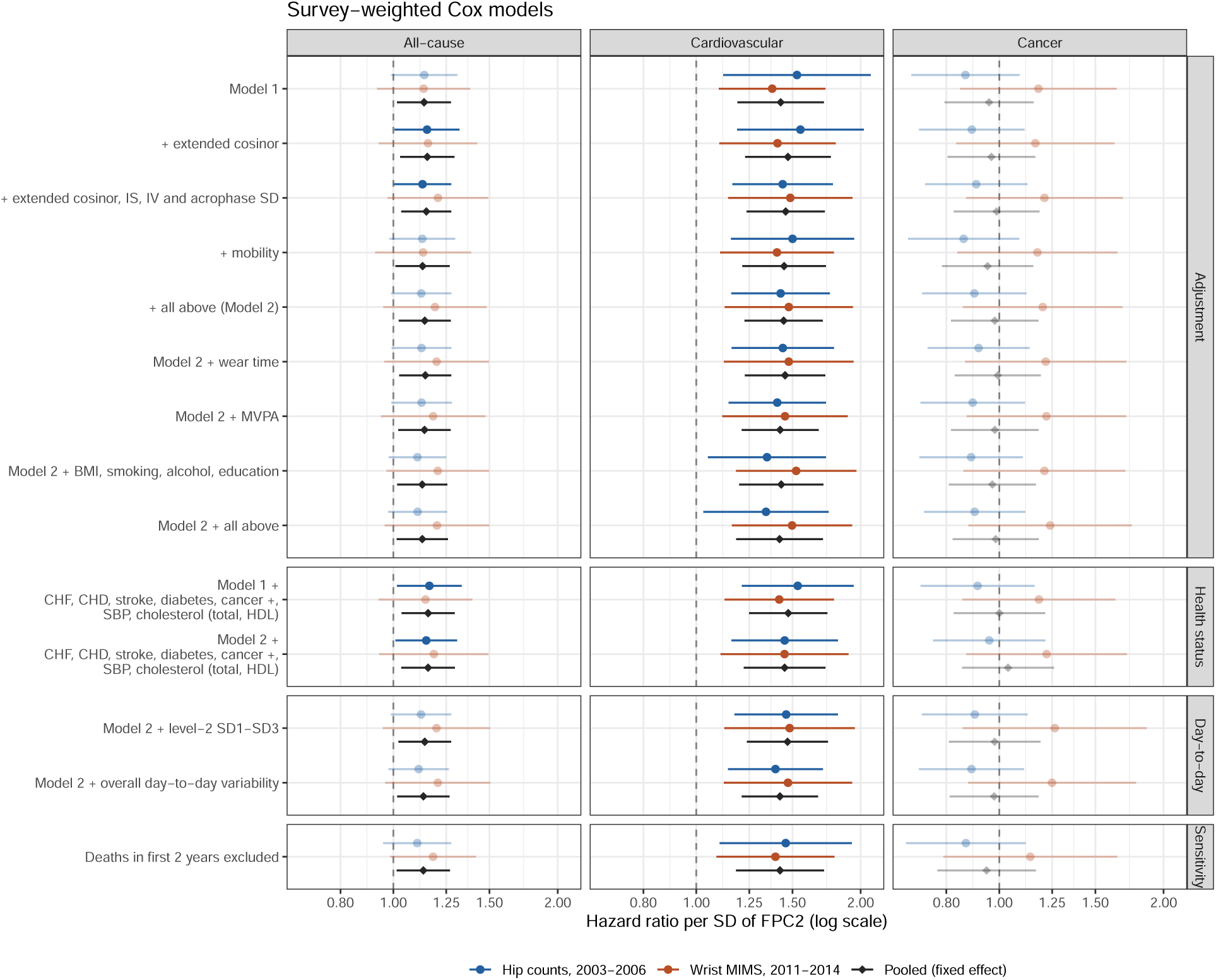
Survey-weighted hazard ratios for all-cause, cardiovascular and cancer mortality per SD of FPC2 under additional adjustments. Model 1: age, sex, ethnicity, income, cosinor mesor, amplitude and acrophase, and FPC1–FPC3. Faded estimates: *p ≥* 0.05.

Adding day-to-day variability from the multilevel FPCA to the models left the association largely unchanged (hazard ratios 1.40 to 1.48). Excluding deaths in the first two years of follow-up gave hazard ratios of 1.46 (95% CI 1.10 to 1.93) and 1.40 (1.09 to 1.79). This argues against reverse causation, in which illness changes activity shortly before death. Results were similar in the expanded sample (Supplementary Tables S2 and S3).

#### 3.2.4 FPC2 and its standard-FPCA counterpart

The identification problems of standard FPCA (Section 3.1.3) also affected its associations with mortality. Without adjustment for the cosinor parameters, the standard counterpart of residual FPC2 (standard FPC4), entered alone with age, sex, ethnicity and income, was not associated with cardiovascular mortality in survey-weighted models (hazard ratio per SD 0.80, 95% CI 0.57 to 1.13, in the hip cohort; 0.96, 0.78 to 1.19, in the wrist cohort); an association emerged only after cosinor adjustment (0.71, 0.51 to 1.00; 0.74, 0.62 to 0.89). Between 23% (hip) and 44% (wrist) of the variance of its scores came from the cosinor part of the average day, which masked the shape signal unless the cosinor parameters were adjusted for. Residual FPC2, which contains no 24-hour component, was associated with cardiovascular mortality with or without this adjustment.

#### 3.2.5 Mortality prediction

In forward selection by cross-validated AUC for cardiovascular death (Figure 7), age was selected first in both cohorts. It was followed by income, mesor and sex in the hip cohort, and by amplitude and income in the wrist cohort. FPC2 was selected next in both cohorts, raising the cross-validated AUC to 0.879 (hip) and 0.890 (wrist); no further variable improved it. FPC2 was also selected by AIC in both cohorts and by EPIC in the wrist cohort, but not by EPIC in the hip cohort. When lifestyle and functional variables were added as candidates, the cross-validated AUC of the selected model increased to 0.896 (hip) and 0.897 (wrist). The gain came mainly from mobility difficulty, which was selected second (hip) or third (wrist), and FPC2 was still selected in both cohorts (Supplementary Figure S10).

**Figure 7:**
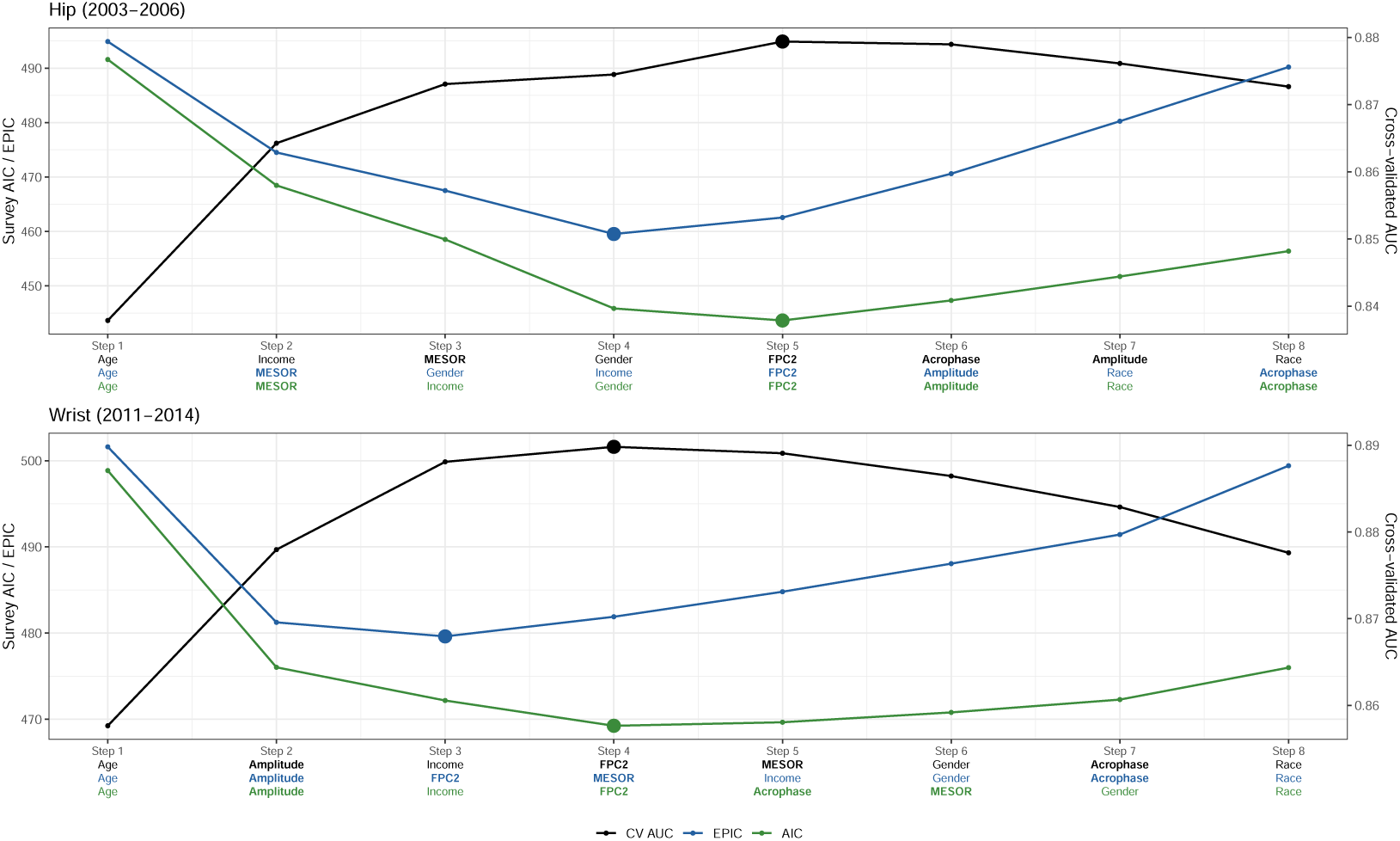
Forward selection of predictors of cardiovascular death. Models were survey-weighted logistic regressions, selected by 10-fold cross-validated AUC, survey AIC or EPIC (AIC with a penalty of 4).

Without lifestyle and functional variables, the final models for predicting cardiovascular death were

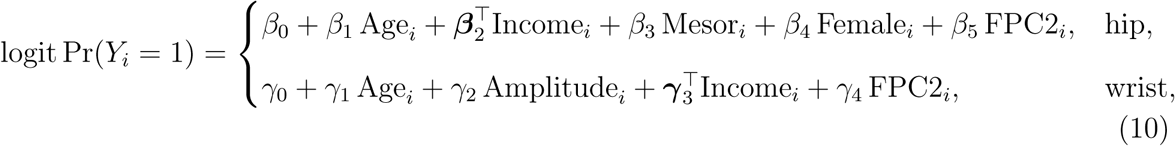

where Income*_i_* is the vector of indicators for the income categories relative to the reference category. The estimated coefficients were

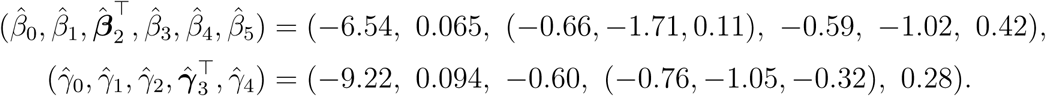

### 3.3 Absence of distinct activity phenotypes

All *k*-means fits converged. The FPCA scores of the activity profiles (13 components explaining 91.6% and 91.8% of variance) did not form well-separated clusters. The gap statistic selected *K* = 3 in the hip and *K* = 4 in the wrist cohort, but the average silhouette width was 0.20 and 0.18 for the selected solutions and at most 0.21 and 0.23 for any *K* from 2 to 8, below the 0.25 threshold for substantial structure throughout (Supplementary Figure S8). In the plane of the first two components, the clusters partitioned a single continuous cloud of points rather than separating distinct groups (Supplementary Figure S9). Activity profiles therefore varied continuously between people, which supports the use of continuous component scores in health analyses. The clusters, and their average 24-hour profiles (Figure 8), are provided in the interactive tool as descriptive groupings along this continuum, not as distinct phenotypes.

**Figure 8:**
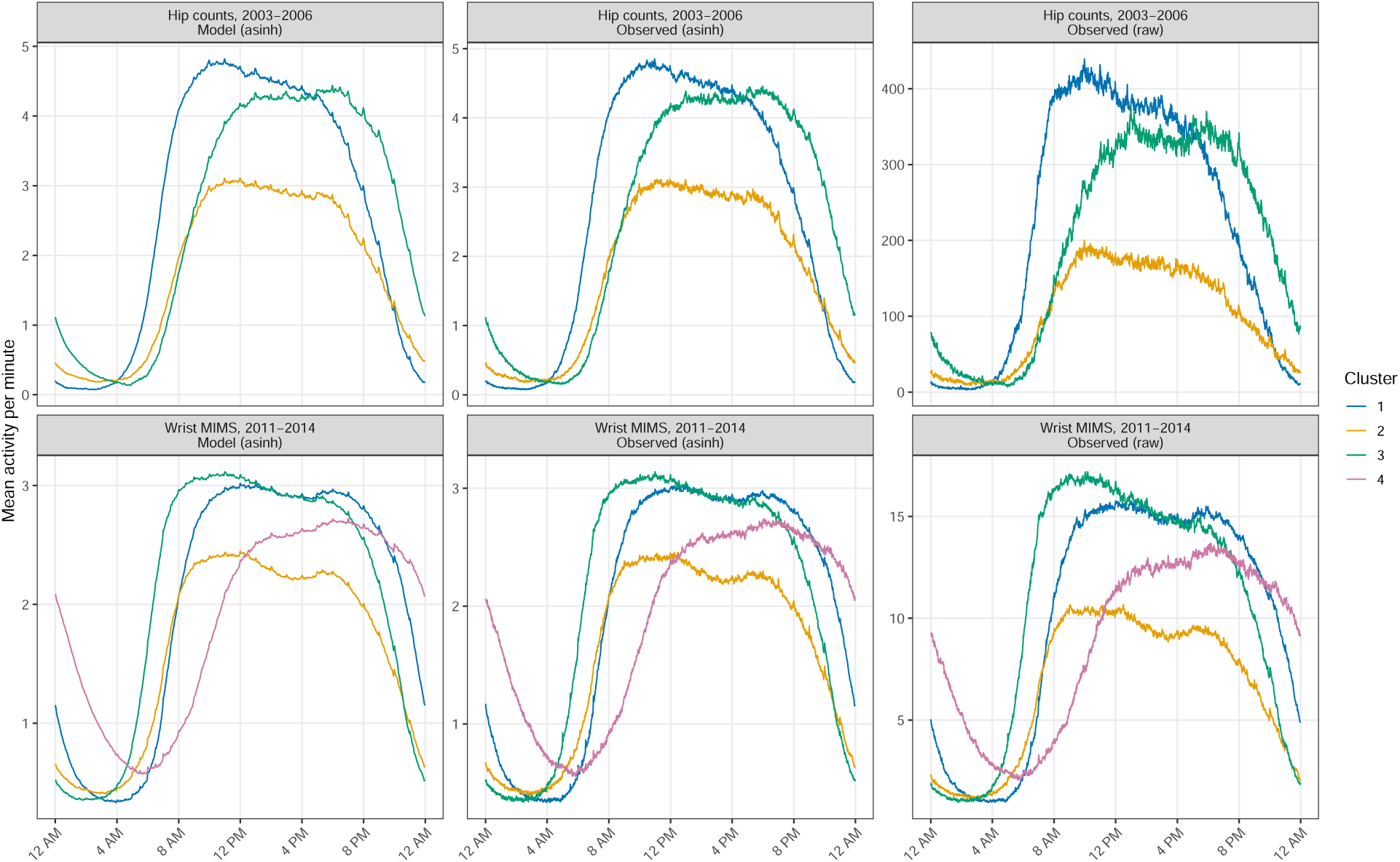
Average 24-hour profile of each cluster: mean of the FPCA-fitted curves (model, asinh scale) and of the observed participant-average curves (asinh and original scale).

### 3.4 Interactive tool

Combined with the cosinor parameters and covariates, the score gives a predicted mortality risk from the final prediction model. We developed a web application (https://quq-tot-orz.shinyapps.io/fpca-residual-tool/) in which users upload minute-level hip activity counts or wrist MIMS units, ideally for seven days. The tool fits the cosinor, computes the residual shape and its FPC2 score from the fixed NHANES reference (mean, eigenfunction and FACE shrinkage factor), and reports the score in cohort SD units and percentiles alongside the mesor, amplitude and acrophase. It also assigns the nearest descriptive activity group and, from the selected logistic model, an estimated probability of cardiovascular death over the NHANES follow-up with its 95% confidence interval. The output is cohort-based context, not an individual diagnosis.

## 4 Discussion

In two NHANES cohorts measured with different devices, we separated each participant’s average day into a 24-hour cosinor rhythm and a residual shape. The leading shape components were 12-hour and 8-hour patterns that the cosinor cannot represent, and activity profiles varied continuously between people rather than forming distinct phenotypes. One shape component, FPC2, which contrasts an early-starting, flat day with a day with distinct morning and evening activity peaks, had the same shape in both cohorts. It was associated with higher all-cause mortality in unweighted analyses in both cohorts. The association was concentrated in cardiovascular deaths, which were associated with FPC2 in both cohorts with survey weights, and it was unchanged after excluding deaths in the first two years and after adjustment for additional rhythm metrics, activity volume, lifestyle and disease factors. FPC2 was not associated with cancer mortality.

Functional data generally contain both phase and amplitude, but traditional methods often ignore the phase part and model only amplitude, using techniques like FPCA [42]. In our study, the decomposition into the cosinor fit and the residual curve is orthogonal, so the two cover non-overlapping parts of the daily profile. Applying FPCA to the residuals yields shape components that do not overlap with the mean activity level, amplitude, and timing; in addition, multilevel FPCA [43] further distinguishes a participant’s usual shape from day-to-day variation. The components of our approach, the cosinor model and functional data analysis, are well established [44]; our contribution lies in combining them so that the added value of the functional representation can be assessed directly and cleanly. Our results answer the question raised in the introduction. Once the cosinor fit is removed, the remaining daily shape is dominated by two peaks that varies in its timing (FPC1) and prominence (FPC2). Neither a single 24-hour cosine nor the extended cosinor can capture such multimodal, asymmetric shapes.

Removing the cosinor before FPCA also changed which patterns were identified and how reliably. In standard FPCA of the average curves, the 24-hour harmonic dominates between-person variance and is correlated with the shorter harmonics, so the two-peak patterns appeared only as lower-ranked components that also carried 24-hour variation, and the nearequal variance of the cosine and sine directions of the 12-hour harmonic left them nearly tied. The low end of residual FPC2 resembles the bi-phasic profile described by Xiao et al. [10]. In samples of a few hundred participants, typical of many accelerometry studies, these components were frequently misplaced or distorted, whereas the residual components were reproduced consistently. The mixing also affected the mortality association: about a quarter (hip) to almost half (wrist) of the variance of the standard counterpart of FPC2 came from the cosinor part of the day, and its association with cardiovascular mortality appeared only after adjustment for the cosinor parameters. This may explain why shape components in standard FPCA have rarely been linked to mortality. In MrOS, Zeitzer et al. [9] described a component contrasting morning and late-afternoon activity similar in shape to FPC2, without adjustment for the cosinor parameters. Residual FPCA makes this adjustment unnecessary, because the components contain no 24-hour variation by construction.

In our models, cosinor amplitude or mesor remained the strongest rhythm predictor of mortality, consistent with previous studies reporting associations between rest–activity amplitude and mortality [23, 24, 25]. In contrast, the 12-hour harmonic component (FPC2) was associated with cardiovascular but not all-cause mortality in the survey-weighted models. A link between this structure and cardiovascular disease is biologically plausible. Disrupted daily rest–activity rhythms were associated with higher mortality, cardiovascular disease and accelerated biological ageing [9, 41, 45], and 12-hour rhythms in gene expression and metabolism are coordinated by a cell-autonomous 12-hour clock distinct from the circadian clock [46]. Cardiovascular physiology also has a 12-hour structure: blood pressure follows a bimodal daily course [47, 48, 49], and an exaggerated morning surge predicts stroke independently of 24-hour blood pressure [50]. The higher-risk end of FPC2 combines more activity at night, an earlier start, no mid-morning surge and less evening activity. This pattern could arise from disrupted sleep or nocturnal activity, or from fewer periods of structured or discretionary activity during the day. The cardiovascular association was not explained by income, mobility difficulty or activity volume, and it persisted after excluding early deaths, which argues against a simple explanation by illness-related inactivity. Our study showed a robust association between the 12-hour component (FPC2) and cardiovascular mortality, suggesting that FPC2 may serve as an early behavioural marker of subclinical cardiovascular disease.

The association between FPC2 and mortality differed between the unweighted and surveyweighted Cox models. The two models answer different questions [51]: the weighted model estimates the association averaged over the U.S. adult population, whereas the unweighted model estimates the association in the sampled cohort, conditional on the covariates included. NHANES oversamples older adults, who therefore account for most deaths while carrying relatively little survey weight. Our results showed that survey weighting moved the all-cause and cardiovascular estimates in opposite directions. These findings should be confirmed in larger cohorts with more deaths, which would also allow age-stratified associations to be examined. In addition, further adjustment for other potential confounders, such as weekday–weekend differences and shift work , could be considered in future analyses of the association between the proposed shape component and mortality.

Several studies have applied functional clustering to identify representative physical behaviour profiles [52, 53, 54]. In these studies, cluster validity was usually assessed by visual separation of cluster trajectories and by differences in health outcomes between clusters [17, 19]. In our data, activity profiles varied continuously between participants, with no clear cluster structure. This is consistent with [19], who reported only small statistical differences between clusters derived from 36 accelerometer metrics covering rest–activity rhythm, daytime activity, sleep and chronotype. We therefore consider continuous component scores the appropriate predictors in health analyses, while clusters remain useful as descriptive profile types for communicating results, as in our web application. The mortality prediction model in the interactive tool asks users for only a few personal details (age, sex and income category) in addition to their accelerometry data. It still achieved a relatively high AUC while limiting the personal information users must share.

This study has several strengths. The decomposition separates rhythm from shape by construction. The analysis was replicated in an independent cohort with a different device placement and measurement unit, and the leading components had nearly identical shapes in both cohorts. The findings were also robust to extensive covariate adjustment. The residual components were more stable than their standard-FPCA counterparts, particularly in small samples, and the association of FPC2 with mortality was independent of adjustment for the cosinor parameters. The study also has limitations. Minutes without valid data were set to zero. This mainly affected the hip cohort at night, when the device was removed, so the hip residual partly reflects device removal rather than activity. In addition, hip counts and wrist MIMS capture different aspects of movement, so components with the same shape should be interpreted with caution across cohorts.

## 5 Conclusion

The within-day structure of activity that a 24-hour cosinor cannot represent varies continuously between people. Its second leading component contrasts a day with distinct morning and evening activity peaks against an early-starting, flat day. This component was associated with higher all-cause or cardiovascular mortality in two independent NHANES cohorts, independently of 24-hour rhythm strength, timing and activity volume, although the specific outcome depended on survey weighting. These exploratory findings suggest that the shape of daily activity carries prognostic information beyond standard rhythm summaries. Such shape components show potential value for personalized medicine, helping to identify individuals at elevated risk and to tailor interventions to their daily activity patterns.

## Data and code availability

NHANES accelerometry, covariate and linked mortality data are publicly available from the National Center for Health Statistics. Analysis code is available at github repo https://github.com/QuQ-ToT-Orz/FPCA_residual, and the interactive tool at https://quq-tot-orz.shinyapps.io/fpca-residual-tool/.

## Authors’ contributions

X.Z. conceived and designed the study, developed the methodology, performed the statistical analyses, and drafted the manuscript.

## Acknowledgments

This project has received funding from the European Union’s Horizon Europe research and innovation programme under the Marie Skłodowska-Curie Actions grant agreement No. 101072993 (Project: LABDA).

## References

[1] Andrew Leroux et al. “Organizing and Analyzing the Activity Data in NHANES”. en. In: Statistics in Biosciences 11.2 (July 2019), pp. 262–287. issn: 1867-1764, 1867-1772. doi: 10.1007/s12561-018-09229-9. url: http://link.springer.com/10.1007/s12561-018-09229-9.

[2] Aiden Doherty et al. “Large Scale Population Assessment of Physical Activity Using Wrist Worn Accelerometers: The UK Biobank Study”. en. In: PLOS ONE 12.2 (Feb. 2017). Ed. by Maciej Buchowski, e0169649. issn: 1932-6203. doi: 10.1371/journal.pone.0169649. url: https://dx.plos.org/10.1371/journal.pone.0169649.

[3] Francesco Sera et al. “Using functional data analysis to understand daily activity levels and patterns in primary school-aged children: Cross-sectional analysis of a UK-wide study”. en. In: PLOS ONE 12.11 (Nov. 2017). Ed. by Rod K. Dishman, e0187677. issn: 1932-6203. doi: 10.1371/journal.pone.0187677. url: https://dx.plos.org/10.1371/journal.pone.0187677.

[4] Jeff Goldsmith et al. “New Insights into Activity Patterns in Children, Found Using Functional Data Analyses”. en. In: Medicine & Science in Sports & Exercise 48.9 (Sept. 2016), pp. 1723–1729. issn: 0195-9131. doi: 10.1249/MSS.0000000000000968. url: https://www.ovid.com/00005768-201609000-00011.

[5] Jan Gertheiss et al. Functional Data Analysis: An Introduction and Recent Developments. en. arXiv:2312.05523 [stat.ME]. Dec. 2023. doi: 10.48550/arXiv.2312.05523. url: http://arxiv.org/abs/2312.05523.

[6] Jia Wang et al. “Measuring the impact of apnea and obesity on circadian activity patterns using functional linear modeling of actigraphy data”. en. In: Journal of Circadian Rhythms 9.0 (Oct. 2011), p. 11. issn: 1740-3391. doi: 10.1186/1740-3391-9-11. url: https://www.jcircadianrhythms.com/article/10.1186/1740-3391-9-11/.

[7] Michael J. Stein et al. “Association between circadian physical activity patterns and mortality in the UK Biobank”. en. In: International Journal of Behavioral Nutrition and Physical Activity 20.1 (Sept. 2023), p. 102. issn: 1479-5868. doi: 10.1186/s12966-023-01508-z. url: https://ijbnpa.biomedcentral.com/articles/10.1186/s12966-023-01508-z.

[8] Sonia Difrancesco et al. “Sociodemographic, Health and Lifestyle, Sampling, and Mental Health Determinants of 24-Hour Motor Activity Patterns: Observational Study”. en. In: Journal of Medical Internet Research 23.2 (Feb. 2021), e20700. issn: 1438-8871. doi: 10.2196/20700. url: http://www.jmir.org/2021/2/e20700/.

[9] Jamie M Zeitzer et al. “Daily Patterns of Accelerometer Activity Predict Changes in Sleep, Cognition, and Mortality in Older Men”. en. In: The Journals of Gerontology: Series A 73.5 (Apr. 2018), pp. 682–687. issn: 1079-5006, 1758-535X. doi: 10.1093/gerona/glw250. url: https://academic.oup.com/biomedgerontology/article/73/5/682/2967602.

[10] Qian Xiao et al. “Rest-activity profiles among U.S. adults in a nationally representative sample: a functional principal component analysis”. en. In: International Journal of Behavioral Nutrition and Physical Activity 19.1 (Mar. 2022), p. 32. issn: 1479-5868. doi: 10.1186/s12966-022-01274-4. url: https://ijbnpa.biomedcentral.com/articles/10.1186/s12966-022-01274-4.

[11] Chris Ho, Ching Yeung et al. “U.S. Adolescent Rest-Activity patterns: insights from functional principal component analysis (NHANES 2011–2014)”. en. In: International Journal of Behavioral Nutrition and Physical Activity 20.1 (Oct. 2023), p. 125. issn: 1479-5868. doi: 10.1186/s12966-023-01520-3. url: https://ijbnpa.biomedcentral.com/articles/10.1186/s12966-023-01520-3.

[12] Chong-Zhi Di et al. “Multilevel functional principal component analysis”. en. In: The Annals of Applied Statistics 3.1 (Mar. 2009), pp. 458–488. issn: 1932-6157, 1941-7330. doi: 10.1214/08-AOAS206. url: https://projecteuclid.org/journals/annals-of-applied-statistics/volume-3/issue-1/Multilevel-functional-principal-component-analysis/10.1214/08-AOAS206.full.

[13] Erjia Cui et al. “Fast Multilevel Functional Principal Component Analysis”. en. In: Journal of Computational and Graphical Statistics 32.2 (Apr. 2023), pp. 366–377. issn: 1061-8600, 1537-2715. doi: 10.1080/10618600.2022.2115500. url: https://www.tandfonline.com/doi/full/10.1080/10618600.2022.2115500.

[14] Xinkai Zhou et al. Generalized Multilevel Functional Principal Component Analysis with Application to NHANES Active Inactive Patterns. en. arXiv:2311.14054 [stat.ME]. Mar. 2025. doi: 10.48550/arXiv.2311.14054. url: http://arxiv.org/abs/2311.14054.

[15] Álvaro Méndez-Civieta et al. “Functional quantile principal component analysis”. en. In: Biostatistics 26.1 (Dec. 2024), kxae040. issn: 1468-4357. doi: 10.1093/biostatistics/kxae040. url: https://academic.oup.com/biostatistics/article/doi/10.1093/biostatistics/kxae040/7831706.

[16] Julien Jacques and Cristian Preda. “Functional data clustering: a survey”. en. In: Advances in Data Analysis and Classification 8.3 (Sept. 2014), pp. 231–255. issn: 1862-5347, 1862-5355. doi: 10.1007/s11634-013-0158-y. url: http://link.springer.com/10.1007/s11634-013-0158-y.

[17] David Lopez-Veneros et al. “Characterization and clustering of intra-day physical activity patterns using accelerometry among sexual and gender minority adults”. en. In: BMC Public Health 25.1 (July 2025), p. 2294. issn: 1471-2458. doi: 10.1186/s12889-025-23425-5. url: https://bmcpublichealth.biomedcentral.com/articles/10.1186/s12889-025-23425-5.

[18] Wookyeong Song et al. Multi-feature Clustering of Step Data using Multivariate Functional Principal Component Analysis. en. arXiv:2010.07462 [stat.ME]. Oct. 2020. doi: 10.48550/arXiv.2010.07462. url: http://arxiv.org/abs/2010.07462.

[19] Sam Vidil et al. “Circadian rhythm profiles derived from accelerometer measures of the sleep-wake cycle in two cohort studies”. en. In: Nature Communications 16.1 (Dec. 2025), p. 11357. issn: 2041-1723. doi: 10.1038/s41467-025-66407-2. url: https://www.nature.com/articles/s41467-025-66407-2.

[20] Chenlu Gao et al. “Approaches for Assessing Circadian Rest-Activity Patterns Using Actigraphy in Cohort and Population-Based Studies”. en. In: Current Sleep Medicine Reports 9.4 (Oct. 2023), pp. 247–256. issn: 2198-6401. doi: 10.1007/s40675-023-00267-4. url: https://link.springer.com/10.1007/s40675-023-00267-4.

[21] Junrui Di et al. “Joint and Individual Representation of Domains of Physical Activity, Sleep, and Circadian Rhythmicity”. en. In: Statistics in Biosciences 11.2 (July 2019), pp. 371–402. issn: 1867-1764, 1867-1772. doi: 10.1007/s12561-019-09236-4. url: http://link.springer.com/10.1007/s12561-019-09236-4.

[22] Jinjoo Shim, Elgar Fleisch, and Filipe Barata. “Circadian rhythm analysis using wearable-based accelerometry as a digital biomarker of aging and healthspan”. en. In: npj Digital Medicine 7.1 (June 2024), p. 146. issn: 2398-6352. doi: 10.1038/s41746-024-01111-x. url: https://www.nature.com/articles/s41746-024-01111-x.

[23] Velarie Y. Ansu-Baidoo et al. “Association between circadian rest-activity rhythm and meal timing: NHANES 2013-2014”. en. In: Sleep Health 12.4 (Aug. 2026), pp. 619–627. issn: 23527218. doi: 10.1016/j.sleh.2026.04.008. url: https://linkinghub.elsevier.com/retrieve/pii/S2352721826000756.

[24] Wendy Wang et al. “Association Between Circadian Rest-Activity Rhythms and Incident Dementia in Older Adults: The Atherosclerosis Risk in Communities Study”. en. In: Neurology 106.2 (Jan. 2026), e214513. issn: 0028-3878, 1526-632X. doi: 10.1212/WNL.0000000000214513. url: https://www.neurology.org/doi/10.1212/WNL.0000000000214513.

[25] Erin I. McDonnell et al. “Registration of 24-hour accelerometric rest-activity profiles and its application to human chronotypes”. en. In: Biological Rhythm Research 53.8 (Aug. 2022), pp. 1299–1319. issn: 0929-1016, 1744-4179. doi: 10.1080/09291016.2021.1929673. url: https://www.tandfonline.com/doi/full/10.1080/09291016.2021.1929673.

[26] Matthew R. Marler et al. “The sigmoidally transformed cosine curve: a mathematical model for circadian rhythms with symmetric non-sinusoidal shapes”. en. In: Statistics in Medicine 25.22 (Nov. 2006), pp. 3893–3904. issn: 0277-6715, 1097-0258. doi: 10.1002/sim.2466. url: https://onlinelibrary.wiley.com/doi/10.1002/sim.2466.

[27] Eus J. W. Van Someren et al. “Bright Light Therapy: Improved Sensitivity to Its Effects on Rest-Activity Rhythms in Alzheimer Patients by Application of Nonparametric Methods”. en. In: Chronobiology International 16.4 (Jan. 1999), pp. 505–518. issn: 0742-0528, 1525-6073. doi: 10.3109/07420529908998724. url: https://www.tandfonline.com/doi/full/10.3109/07420529908998724.

[28] Congyu Wu et al. “circadian rhythms are not captured equal: Exploring Circadian metrics extracted by differentcomputational methods from smartphone accelerometer and GPS sensors in daily life tracking”. en. In: DIGITAL HEALTH 8 (Jan. 2022), p. 205520762211142. issn: 2055-2076, 2055-2076. doi: 10.1177/20552076221114201. url: http://journals.sagepub.com/doi/10.1177/20552076221114201.

[29] L. Xiao et al. “Quantifying the lifetime circadian rhythm of physical activity: a covariate-dependent functional approach”. en. In: Biostatistics 16.2 (Apr. 2015), pp. 352–367. issn: 1465-4644, 1468-4357. doi: 10.1093/biostatistics/kxu045. url: https://academic.oup.com/biostatistics/article-lookup/doi/10.1093/biostatistics/kxu045.

[30] Jinjoo Shim and Jukka-Pekka Onnela. “Longitudinal digital phenotyping of activity rhythms and biological aging using commercial wearables”. en. In: Nature Communications 17.1 (Aug. 2026), p. 10155. issn: 2041-1723. doi: 10.1038/s41467-026-76147-6. url: https://www.nature.com/articles/s41467-026-76147-6.

[31] Julia Wrobel et al. “Registration for Exponential Family Functional Data”. en. In: Biometrics 75.1 (Mar. 2019), pp. 48–57. issn: 0006-341X, 1541-0420. doi: 10.1111/biom.12963. url: https://academic.oup.com/biometrics/article/75/1/48-57/7517255.

[32] Dinesh John et al. “An Open-Source Monitor-Independent Movement Summary for Accelerometer Data Processing”. en. In: Journal for the Measurement of Physical Behaviour 2.4 (Dec. 2019), pp. 268–281. issn: 2575-6605, 2575-6613. doi: 10.1123/jmpb.2018-0068. url: https://journals.humankinetics.com/view/journals/jmpb/2/4/article-p268.xml.

[33] Xin Zheng, Ian Meneghel Danilevicz, and Maichina Paw. “Scalable estimation of temporal clustering in accelerometry: a kernel-independent dispersion index grounded in the Hawkes process”. en. In: ().

[34] Marta Karas et al. Comparison of Accelerometry-based Measures of Physical Activity. Mar. 2022. doi:10.1101/2022.03.16.22272518. url: http://medrxiv.org/lookup/doi/10.1101/2022.03.16.22272518.

[35] Luo Xiao et al. “Fast covariance estimation for high-dimensional functional data”. en. In: Statistics and Computing 26.1-2 (Jan. 2016), pp. 409–421. issn: 0960-3174, 1573-1375. doi: 10.1007/s11222-014-9485-x. url: http://link.springer.com/10.1007/s11222-014-9485-x.

[36] Thomas Lumley. “Analysis of Complex Survey Samples”. en. In: Journal of Statistical Software 9.8 (2004). issn: 1548-7660. doi: 10.18637/jss.v009.i08. url: http://www.jstatsoft.org/v09/i08/.

[37] Gary Solon, Steven J. Haider, and Jeffrey M. Wooldridge. What Are We Weighting For? NBER Working Paper 18859. National Bureau of Economic Research, Feb. 2013. url: https://ssrn.com/abstract=2226847.

[38] Nancy R. Cook. “Use and Misuse of the Receiver Operating Characteristic Curve in Risk Prediction”. en. In: Circulation 115.7 (Feb. 2007), pp. 928–935. issn: 0009-7322, 1524-4539. doi: 10.1161/CIRCULATIONAHA.106.672402. url: https://www.ahajournals.org/doi/10.1161/CIRCULATIONAHA.106.672402.

[39] James H. Ware. “The Limitations of Risk Factors as Prognostic Tools”. en. In: New England Journal of Medicine 355.25 (Dec. 2006), pp. 2615–2617. issn: 0028-4793, 1533-4406. doi: 10.1056/NEJMp068249. url: http://www.nejm.org/doi/abs/10.1056/NEJMp068249.

[40] Ekaterina Smirnova et al. “The Predictive Performance of Objective Measures of Physical Activity Derived From Accelerometry Data for 5-Year All-Cause Mortality in Older Adults: National Health and Nutritional Examination Survey 2003–2006”. en. In: The Journals of Gerontology: Series A 75.9 (Sept. 2020). Ed. by Anne Newman,pp. 1779–1785. issn: 1079-5006, 1758-535X. doi: 10.1093/gerona/glz193. url: https://academic.oup.com/biomedgerontology/article/75/9/1779/5555985.

[41] Jinjoo Shim et al. “From wrist data to lifespan: elucidating inflammation-driven biological aging via activity rhythms captured by wearable devices”. en. In: npj Aging 12.1 (Feb. 2026), p. 49. issn: 2731-6068. doi: 10.1038/s41514-026-00349-x. url: https://www.nature.com/articles/s41514-026-00349-x.

[42] J. Derek Tucker, Wei Wu, and Anuj Srivastava. “Generative Models for Functional Data using Phase and Amplitude Separation”. en. In: Computational Statistics & Data Analysis 61 (May 2013). arXiv:1212.1791 [stat.CO], pp. 50–66. issn: 01679473. doi: 10.1016/j.csda.2012.12.001. url: http://arxiv.org/abs/1212.1791.

[43] Selene Yue Xu et al. “Modeling Temporal Variation in Physical Activity Using Functional Principal Components Analysis”. en. In: Statistics in Biosciences 11.2 (July 2019), pp. 403–421. issn: 1867-1764, 1867-1772. doi: 10.1007/s12561-019-09237-3. url: http://link.springer.com/10.1007/s12561-019-09237-3.

[44] Sophie Dabo-Niang and Camille Frévent. Uncovering Data Across Continua: An Introduction to Functional Data Analysis. en. arXiv:2404.16598 [math.ST]. Apr. 2024. doi: 10.48550/arXiv.2404.16598. url: http://arxiv.org/abs/2404.16598.

[45] Lili Huang et al. “Longitudinal association between accelerometer-derived rest–activity rhythm and atherosclerotic cardiovascular disease”. en. In: Sleep Medicine 121 (Sept. 2024), pp. 8–14. issn: 13899457. doi: 10.1016/j.sleep.2024.06.004. url: https://linkinghub.elsevier.com/retrieve/pii/S1389945724002727.

[46] Bokai Zhu et al. “A Cell-Autonomous Mammalian 12 hr Clock Coordinates Metabolic and Stress Rhythms”. en. In: Cell Metabolism 25.6 (June 2017), 1305–1319.e9. issn: 15504131. doi: 10.1016/j.cmet.2017.05.004. url: https://linkinghub.elsevier.com/retrieve/pii/S1550413117302905.

[47] Kuniaki Otsuka et al. “Appropriate Circadian-Circasemidian Coupling Protects Blood Pressure from Morning Surge and Promotes Human Resilience and Wellbeing”. en. In: Clinical Interventions in Aging Volume 18 (May 2023), pp. 755–769. issn: 1178-1998. doi: 10.2147/CIA.S398957. url: https://www.dovepress.com/appropriate-circadian-circasemidian-coupling-protects-blood-pressure-f-peer-reviewed-fulltext-article-CIA.

[48] Yibo Guo et al. “Circadian rhythm pattern of symptom onset in patients with ST-segment elevation myocardial infarction in the Chinese population”. en. In: Frontiers in Cardiovascular Medicine 11 (Dec. 2024), p. 1393390. issn: 2297-055X. doi: 10.3389/fcvm.2024.1393390. url: https://www.frontiersin.org/articles/10.3389/fcvm.2024.1393390/full.

[49] Shougo Murakami, Kuniaki Otsuka, and Tatsuji Kono. “Repeated ambulatory monitoring reveals an evening rise in blood pressure in a Japanese population”. en. In: The Journal of Clinical Hypertension 21.11 (Nov. 2019), pp. 1675–1681. issn: 1524-6175, 1751-7176. doi: 10.1111/jch.13709. url: https://onlinelibrary.wiley.com/doi/10.1111/jch.13709.

[50] Kazuomi Kario et al. “Morning Surge in Blood Pressure as a Predictor of Silent and Clinical Cerebrovascular Disease in Elderly Hypertensives: A Prospective Study”. en. In: Circulation 107.10 (Mar. 2003), pp. 1401–1406. issn: 0009-7322, 1524-4539. doi: 10.1161/01.CIR.0000056521.67546.AA. url: https://www.ahajournals.org/doi/10.1161/01.CIR.0000056521.67546.AA.

[51] Marcos Matabuena and Alexander Petersen. Distributional data analysis of accelerometer data from the NHANES database using nonparametric survey regression models. en. arXiv:2104.01165 [stat.ME]. Jan. 2022. doi:10.48550/arXiv.2104.01165. url: http://arxiv.org/abs/2104.01165.

[52] Vahid Farrahi and Esmaeil Farhang. “Data-Driven Profiling of Accelerometer-Determined Physical Behaviors and Health Outcomes in Adults: A Systematic Review”. In: Journal for the Measurement of Physical Behaviour 9.1 (Jan. 2026), jmpb.2025–0054. issn: 2575-6605, 2575-6613. doi: 10.1123/jmpb.2025-0054. url: https://journals.humankinetics.com/view/journals/jmpb/9/1/article-jmpb.2025-0054.xml.

[53] Jinjoo Shim, Elgar Fleisch, and Filipe Barata. “Wearable-based accelerometer activity profile as digital biomarker of inflammation, biological age, and mortality using hierarchical clustering analysis in NHANES 2011–2014”. en. In: Scientific Reports 13.1 (June 2023), p. 9326. issn: 2045-2322. doi: 10.1038/s41598-023-36062-y. url: https://www.nature.com/articles/s41598-023-36062-y.

[54] Marcos Matabuena et al. “Physical activity phenotypes and mortality in older adults: a novel distributional data analysis of accelerometry in the NHANES”. en. In: Aging Clinical and Experimental Research 34.12 (Oct. 2022), pp. 3107–3114. issn: 1720-8319. doi: 10.1007/s40520-022-02260-3. url: https://link.springer.com/10.1007/s40520-022-02260-3.

