## Supplementary material for "Beyond the 24-Hour Rhythm: Functional Principal Components of Daily Activity Residuals and Mortality in NHANES": supp1

### Supplementary Material for Sensitivity Analyses: study sample, multilevel fPCA, early death exclusion, mortality prediction

Beyond the 24-Hour Rhythm: Functional Principal Components of Daily Activity Residuals and Mortality in NHANES

Xin Zheng*[1,2]

[1]Amsterdam UMC location Vrije Universiteit Amsterdam, Public and Occupational Health, De Boelelaan 1117, Amsterdam, the Netherlands
[2]Amsterdam Public Health, Health Behaviors and Chronic Diseases and Methodology, Amsterdam, the Netherlands

Section S1 (properties of the cosinor-residual decomposition) is provided separately. Unless stated otherwise, hazard ratios (HR) are per cohort SD with 95% confidence intervals; "unweighted" denotes Cox models with equal weights and design-based standard errors accounting for the NHANES strata and primary sampling units, and "survey-weighted" denotes MEC examination weights multiplied by the inverse probability of inclusion in the analytic sample. Model 1 includes age, sex, ethnicity, income, the cosinor mesor, amplitude and acrophase (cosine and sine), and FPC1–FPC3. Model 2 additionally includes the extended cosinor minimum, $\alpha$ and $\log \beta$, interdaily stability (IS), intradaily variability (IV), the circular SD of the daily acrophases and mobility difficulty. Bold: $p < 0.05$. Pooled estimates are fixed-effect inverse-variance averages of the two cohorts.

#### S2.1 Study sample

Table S1 lists the number of participants meeting each exclusion criterion. The main analysis applied all criteria; the expanded sample applied only the accelerometry and mortality-linkage criteria, so that it also includes participants without BMI, education or laboratory

measures and survivors with less than five years of follow-up. The expanded samples comprised 2,917 (hip; 507 deaths, 120 cardiovascular, 109 cancer) and 3,605 participants (wrist; 362, 125 and 91 deaths), aged 18 years or older.

Table S1: Participants meeting each exclusion criterion.

| | Hip, 2003–2006 | Wrist, 2011–2014 |
|---|---|---|
| Participants with accelerometry | 14,631 | 6,905 |
| Age missing (85 years or older in 2003–2006) | 300 | 0 |
| Fewer than 7 valid days[a] | 10,437 | 2,554 |
| Not eligible for, or missing, mortality linkage | 5,029 | 1,318 |
| Missing education[b] | 6,001 | 1,564 |
| Missing body mass index | 96 | 82 |
| Missing systolic blood pressure, total or HDL cholesterol | 1,976 | 932 |
| Alive with less than 5 years of follow-up | 0 | 18 |
| Main analytic sample | 2,615 | 3,208 |
| Expanded sample (valid days and mortality linkage only) | 2,917 | 3,605 |

#### S2.2 Cosinor residual diagnostics

Figures S2 and S3 show normal Q–Q plots and autocorrelation functions of the minute-level residuals of the weekly cosinor fit for the two example participants of Figure S1. In every day, residuals were strongly autocorrelated and heavier-tailed than normal, indicating systematic within-day structure that the cosinor does not capture. Figure S4 shows the average residual of the same participants and its reconstruction from the single-level and level-1 multilevel fPCA.

#### S2.3 Multilevel fPCA

Figures SS5 and Figures SS6 show the multilevel fPCA of the daily residuals. Most residual variance was within participants: the between-participant (level-1, Figures SS5) part accounted for 12.0% (hip) and 14.9% (wrist). The level-2 components (Figures SS6) describe day-to-day deviations, which by construction can include day-to-day changes in level, amplitude and timing (Section S1.8).

#### S2.4 Correlations between rhythm, shape and activity metrics

Spearman correlations among the variables of the mortality models show that total activity, MVPA and wear time were highly correlated with each other ($\rho$ 0.88 to 0.97) and with the

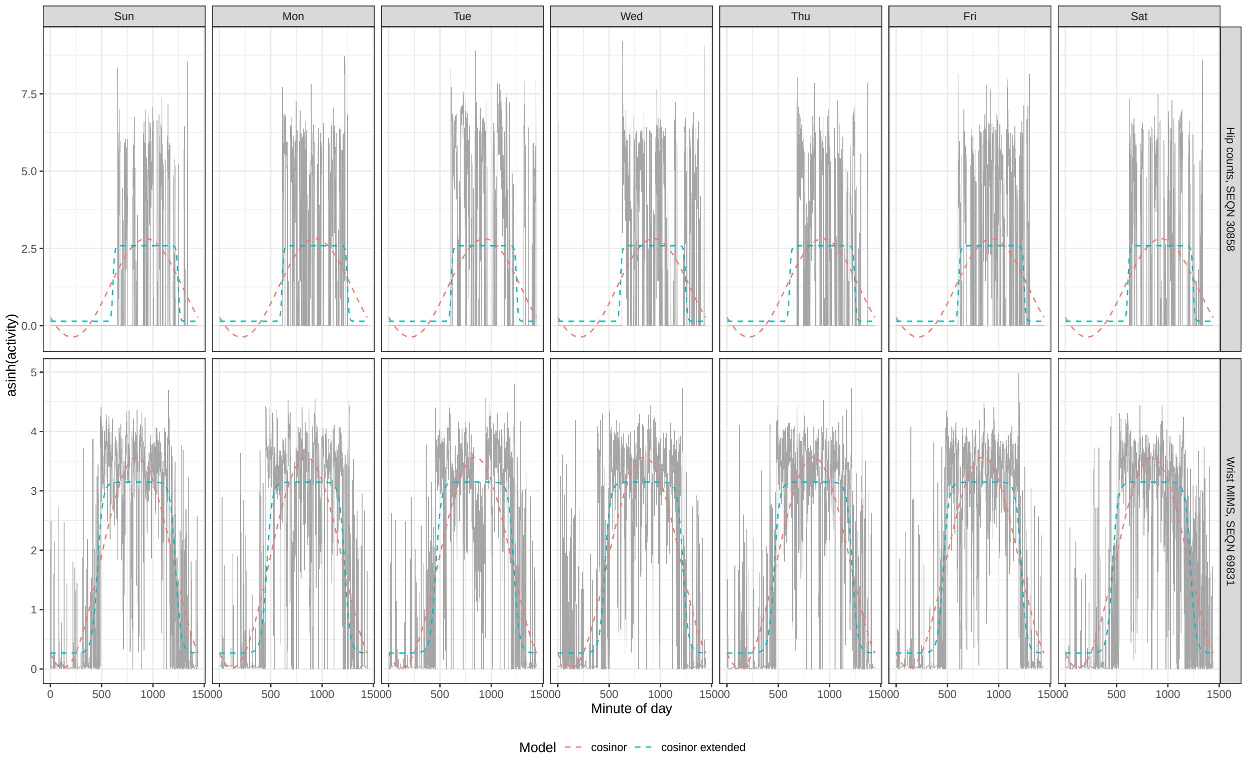


(a)

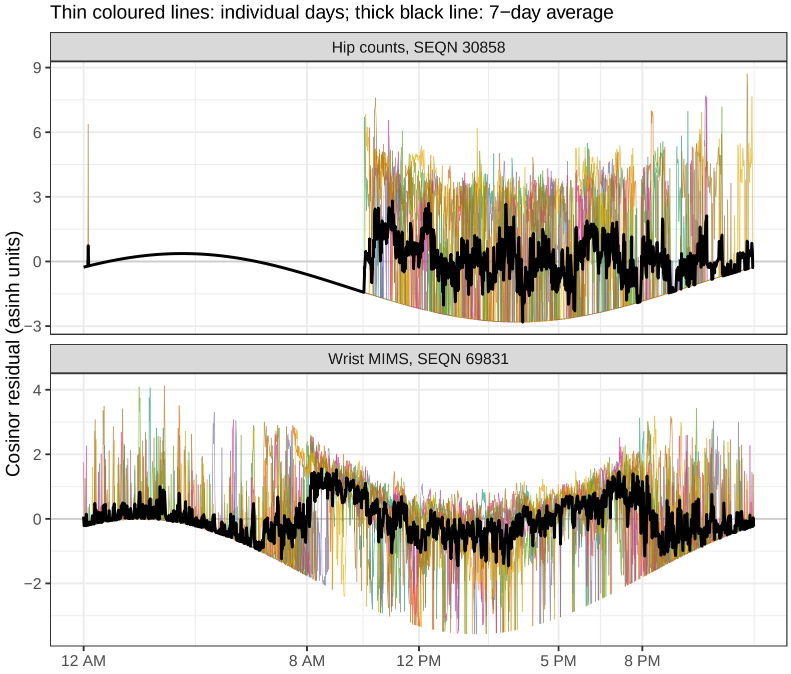


(b)

Figure S1: Decomposition for one hip and one wrist participant. (a) Seven days of asinh-transformed activity (grey) with the participant's 7-day cosinor and extended cosinor fits (dashed). (b) The residual of each day (thin coloured lines) and their average (black), the shape curve analysed by fPCA.

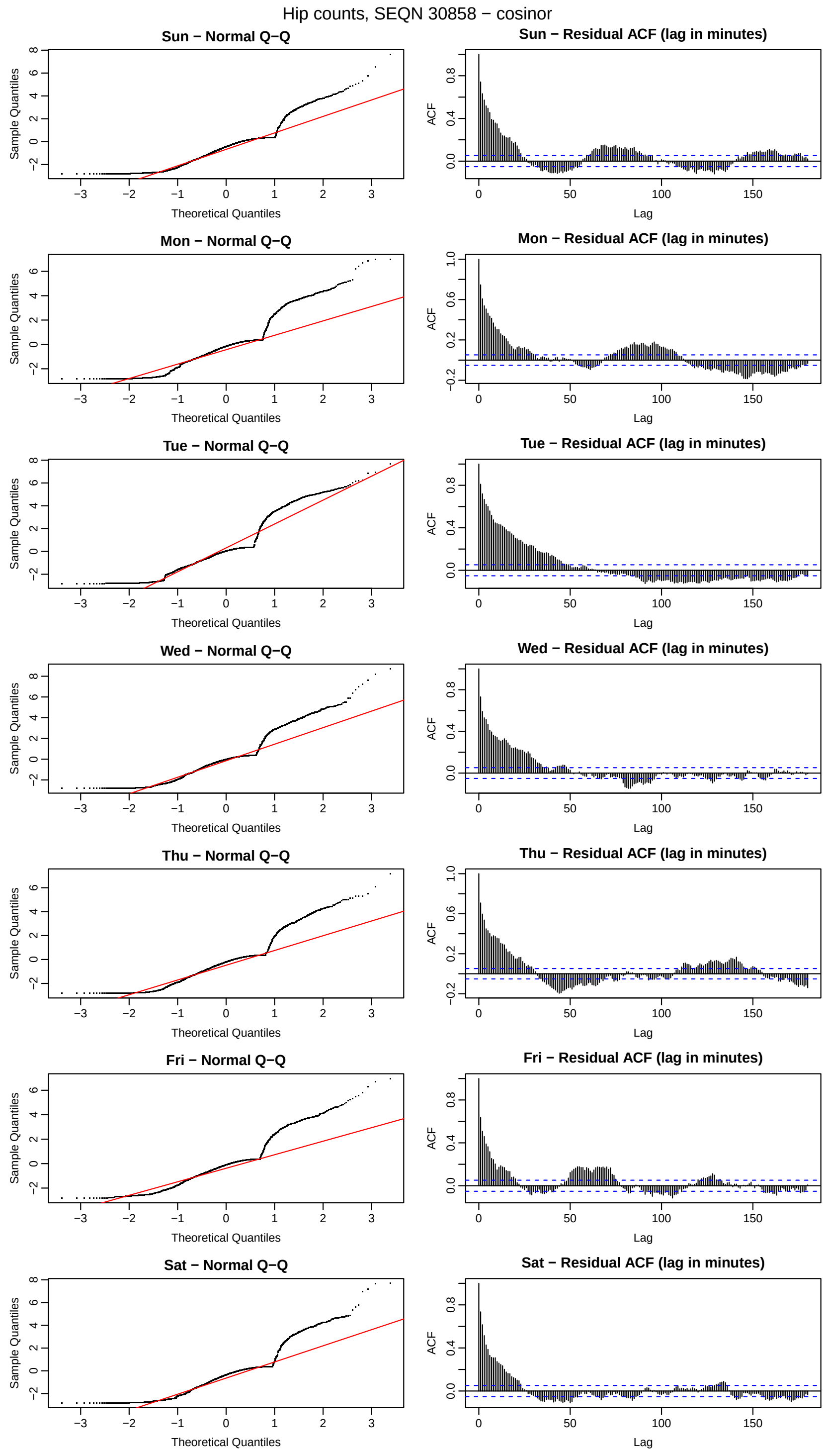


Figure S2: Residual diagnostics for the example hip participant (SEQN 30858): normal Q–Q plot (left) and autocorrelation function up to 180 minutes (right) of the cosinor residual for each day.

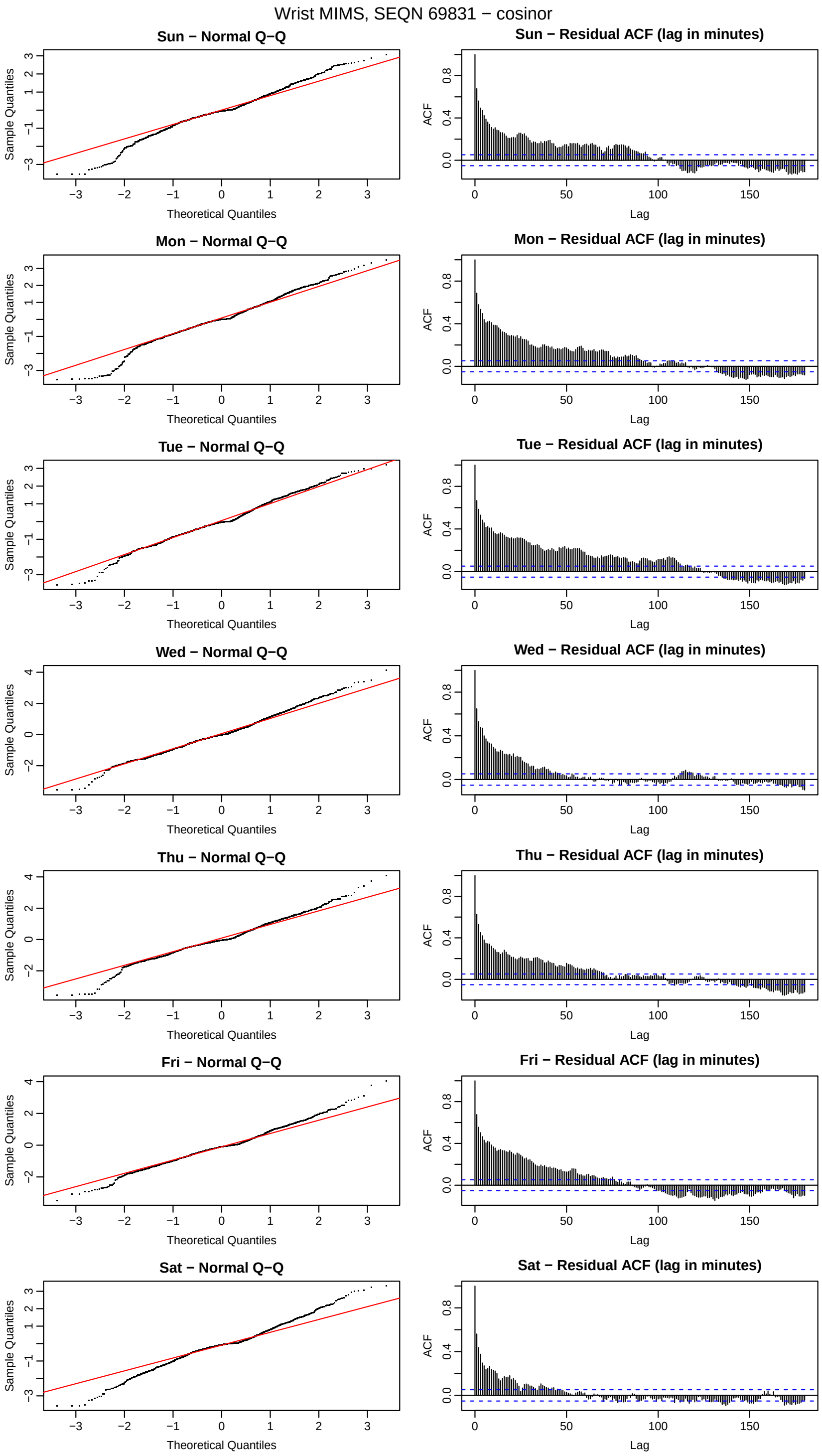


Figure S3: Residual diagnostics for the example wrist participant (SEQN 69831).

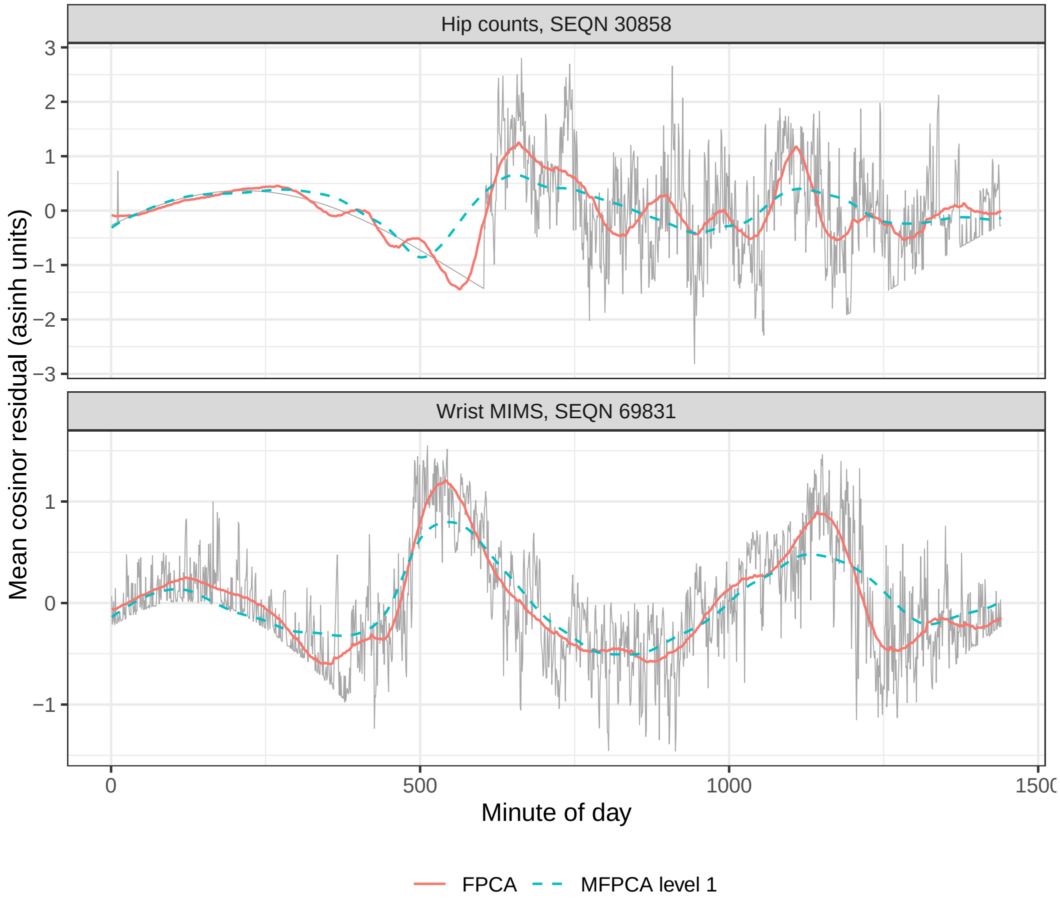


Figure S4: Average cosinor residual of the two example participants (grey) and its reconstruction from the single-level fPCA (solid) and the level-1 component of the multilevel fPCA (dashed).

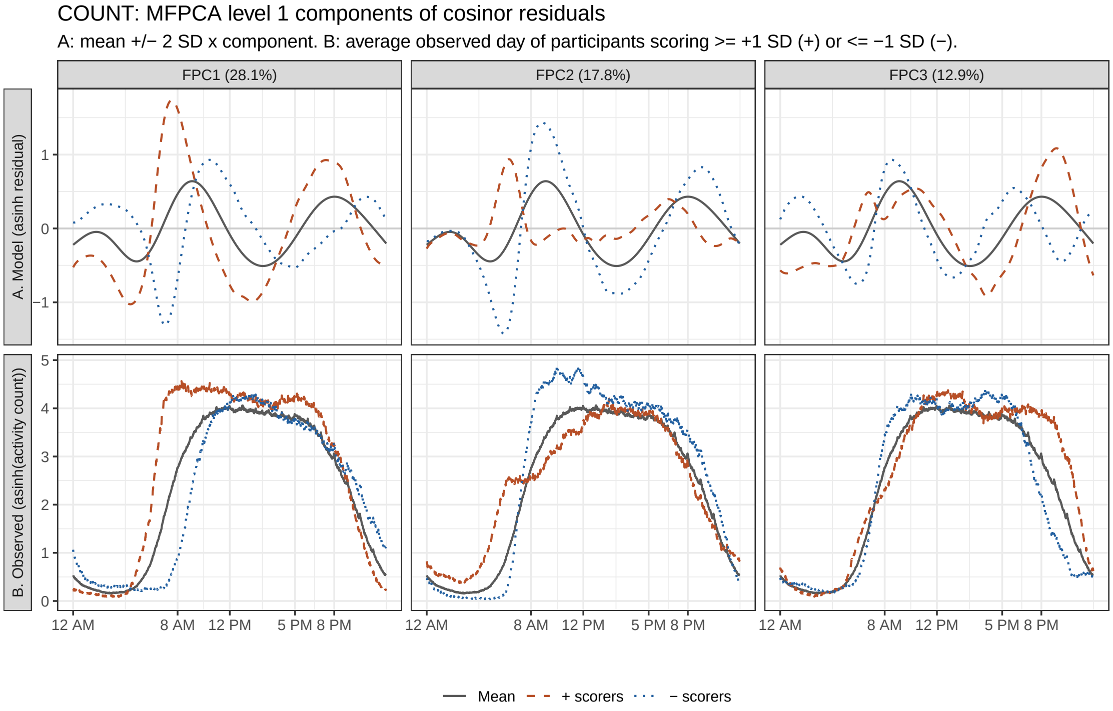


(a) Hip

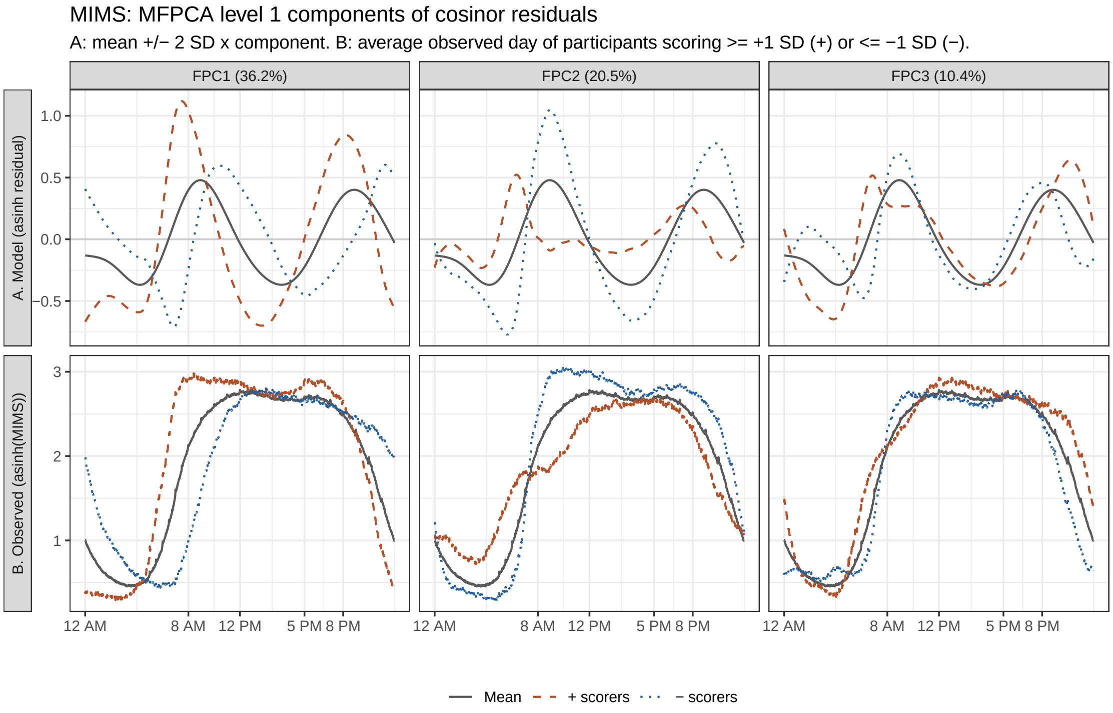


(b) Wrist

Figure S5: Leading level-1 (between-participant) components of the multilevel fPCA.

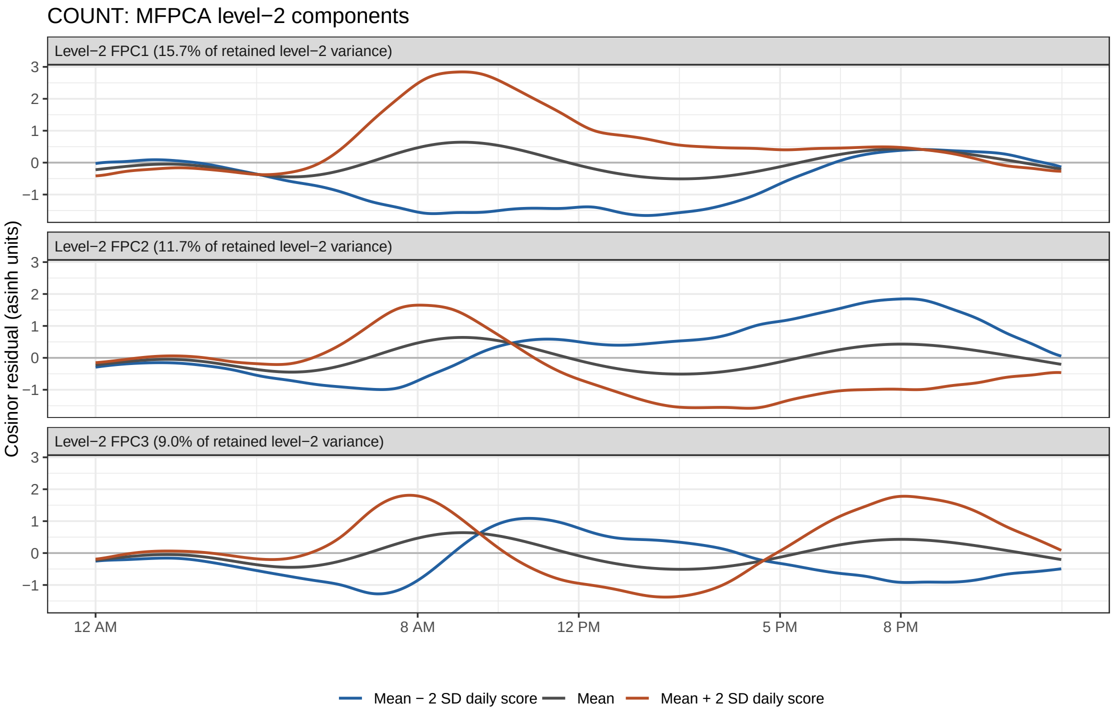


(a) Hip

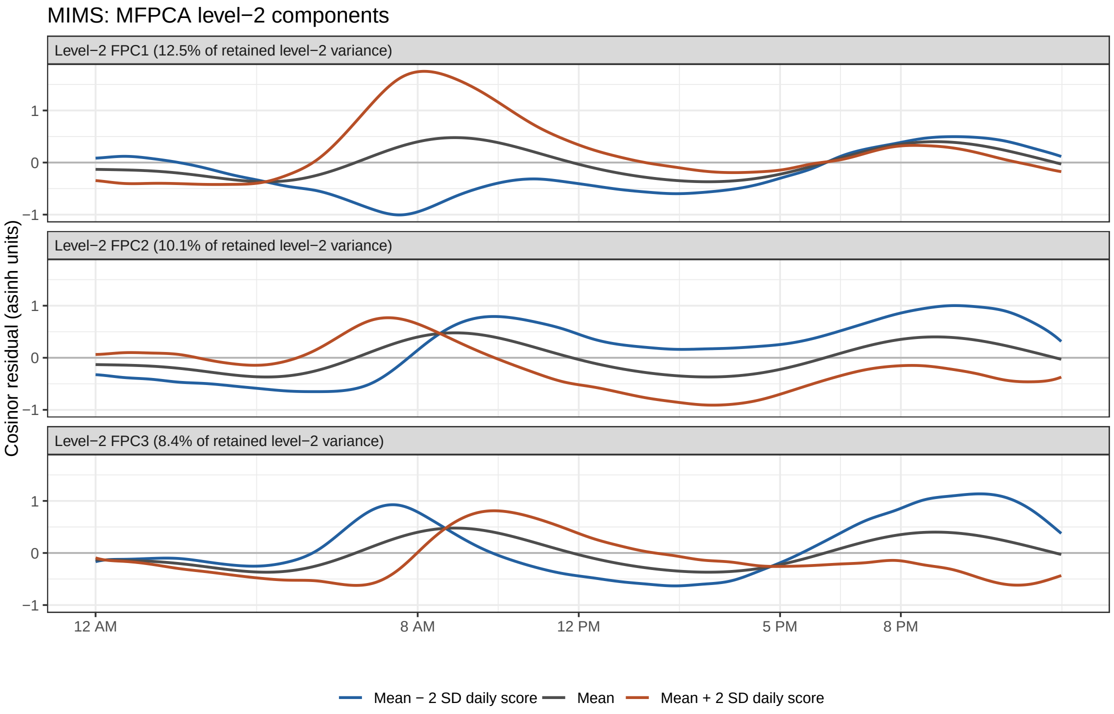


(b) Wrist

Figure S6: Leading level-2 (within-participant, day-to-day) components of the multilevel fPCA.

cosinor mesor ($\rho$ up to 0.93); they were therefore added to Model 2 one at a time and not to Model 1. Supplementary Figure S7 adds the multilevel day-to-day variability summaries.

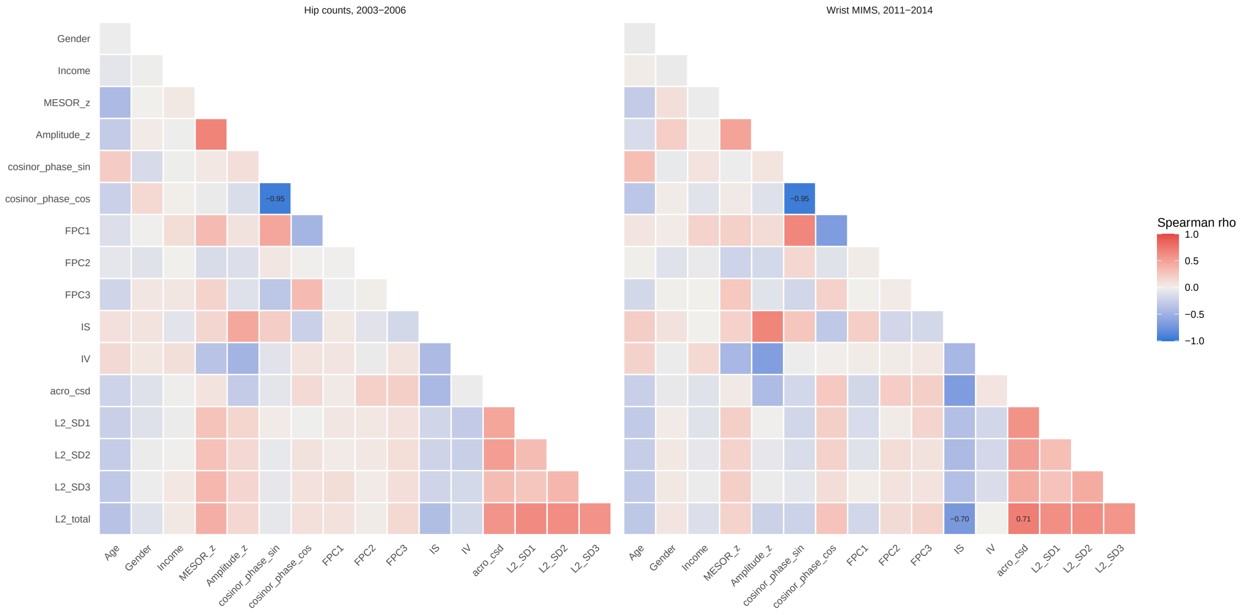


Figure S7: Spearman correlations of the Model 1 variables, IS, IV and the circular SD of the acrophase with the day-to-day variability summaries of the multilevel fPCA (SDs of the three leading level-2 scores and overall day-to-day variability).

#### S2.5 Expanded sample

In the expanded sample, the analysis sequence led to the same conclusions (Table SS2 and Table SS3): FPC2 was the only component associated with all-cause mortality in the discovery cohort ($q = 0.007$) and was replicated in the wrist cohort, and the survey-weighted cardiovascular estimates were 1.53 (1.08 to 2.19) and 1.51 (1.24 to 1.85).

#### S2.6 Clustering diagnostics

Clustering used the fPCA scores of the participant-average asinh activity curves (13 components in each cohort, explaining 91.6% and 91.8% of variance). Figure S9 gives the gap statistic and average silhouette width for $K = 1, \ldots, 8$, which did not reach 0.25 for any $K$. Figure S8 shows that the FPC1 and FPC2 scores form a single continuous cloud.

Table S2: Model 1 hazard ratios per SD of the shape components, survey-weighted (inverse-probability-of-inclusion weights), expanded sample.

| | Hip counts, 2003–2006 | | | Wrist MIMS, 2011–2014 | | | Pooled | | |
|---|---|---|---|---|---|---|---|---|---|
| | HR (95% CI) | $p$ | PH $p$ | HR (95% CI) | $p$ | PH $p$ | HR (95% CI) | $p$ | Het. $p$ |
| *All-cause mortality* | | | | | | | | | |
| Deaths / $n$ | 507 / 2,917 | | | 362 / 3,605 | | | 869 / 6,522 | | |
| FPC1 | 0.99 (0.80–1.22) | 0.911 ($q$=0.911) | 0.870 | 0.91 (0.76–1.10) | 0.332 | 0.284 | | | |
| FPC2 | 1.12 (0.97–1.30) | 0.123 ($q$=0.370) | 0.701 | 1.13 (0.95–1.36) | 0.175 | 0.953 | **1.13 (1.01–1.26)** | **0.040** | 0.935 |
| FPC3 | 0.94 (0.82–1.07) | 0.332 ($q$=0.499) | 0.176 | 0.97 (0.83–1.14) | 0.742 | 0.135 | | | |
| *Cardiovascular mortality* | | | | | | | | | |
| Deaths / $n$ | 120 / 2,917 | | | 125 / 3,605 | | | 245 / 6,522 | | |
| FPC1 | 1.11 (0.85–1.44) | 0.439 | 0.281 | 0.82 (0.62–1.10) | 0.190 | 0.834 | | | |
| FPC2 | **1.53 (1.08–2.19)** | **0.018** | 0.847 | **1.51 (1.24–1.85)** | **<0.001** | 0.661 | **1.52 (1.28–1.81)** | **<0.001** | 0.949 |
| FPC3 | **0.68 (0.50–0.93)** | **0.014** | 0.589 | 1.01 (0.77–1.32) | 0.940 | 0.738 | | | |
| *Cancer mortality* | | | | | | | | | |
| Deaths / $n$ | 109 / 2,917 | | | 91 / 3,605 | | | 200 / 6,522 | | |
| FPC1 | 1.21 (0.82–1.79) | 0.326 | 0.757 | 1.01 (0.74–1.40) | 0.929 | 0.093 | | | |
| FPC2 | 0.80 (0.63–1.01) | 0.064 | 0.433 | 1.09 (0.80–1.49) | 0.570 | 0.746 | 0.90 (0.74–1.08) | 0.259 | 0.115 |
| FPC3 | 1.15 (0.76–1.75) | 0.503 | 0.982 | 1.04 (0.69–1.55) | 0.860 | 0.212 | | | |

Models adjusted for age, sex, ethnicity, income, cosinor mesor, amplitude and acrophase. $q$: Benjamini–Hochberg adjusted over FPC1–FPC3 in the hip discovery cohort for all-cause mortality. PH $p$: Schoenfeld residual test. Pooled: fixed-effect inverse-variance estimate for FPC2; Het. $p$: Cochran's $Q$ test. Bold: $p < 0.05$.

Table S3: Model 1 hazard ratios per SD of the shape components, unweighted, with design-based standard errors, expanded sample.

| | Hip counts, 2003–2006 | | | Wrist MIMS, 2011–2014 | | | Pooled | | |
|---|---|---|---|---|---|---|---|---|---|
| | HR (95% CI) | $p$ | PH $p$ | HR (95% CI) | $p$ | PH $p$ | HR (95% CI) | $p$ | Het. $p$ |
| *All-cause mortality* | | | | | | | | | |
| Deaths / $n$ | 507 / 2,917 | | | 362 / 3,605 | | | 869 / 6,522 | | |
| FPC1 | 1.05 (0.90–1.23) | 0.525 ($q$=0.525) | 0.870 | 0.96 (0.83–1.09) | 0.513 | 0.284 | | | |
| FPC2 | **1.16 (1.05–1.27)** | **0.002 ($q$=0.007)** | 0.701 | **1.20 (1.05–1.36)** | **0.007** | 0.953 | **1.17 (1.09–1.26)** | **<0.001** | 0.685 |
| FPC3 | 1.05 (0.94–1.17) | 0.372 ($q$=0.525) | 0.176 | 0.98 (0.86–1.11) | 0.700 | 0.135 | | | |
| *Cardiovascular mortality* | | | | | | | | | |
| Deaths / $n$ | 120 / 2,917 | | | 125 / 3,605 | | | 245 / 6,522 | | |
| FPC1 | 1.07 (0.86–1.35) | 0.535 | 0.281 | 0.97 (0.76–1.23) | 0.778 | 0.834 | | | |
| FPC2 | 1.22 (0.94–1.57) | 0.135 | 0.847 | **1.40 (1.18–1.66)** | **<0.001** | 0.661 | **1.34 (1.17–1.55)** | **<0.001** | 0.368 |
| FPC3 | 0.89 (0.68–1.17) | 0.405 | 0.589 | 0.93 (0.75–1.17) | 0.555 | 0.738 | | | |
| *Cancer mortality* | | | | | | | | | |
| Deaths / $n$ | 109 / 2,917 | | | 91 / 3,605 | | | 200 / 6,522 | | |
| FPC1 | 1.08 (0.83–1.40) | 0.557 | 0.757 | 1.13 (0.83–1.53) | 0.443 | 0.093 | | | |
| FPC2 | 0.92 (0.77–1.09) | 0.320 | 0.433 | 1.08 (0.81–1.44) | 0.590 | 0.746 | 0.96 (0.83–1.11) | 0.563 | 0.331 |
| FPC3 | 1.12 (0.85–1.49) | 0.420 | 0.982 | 1.12 (0.86–1.47) | 0.392 | 0.212 | | | |

Models adjusted for age, sex, ethnicity, income, cosinor mesor, amplitude and acrophase. $q$: Benjamini–Hochberg adjusted over FPC1–FPC3 in the hip discovery cohort for all-cause mortality. PH $p$: Schoenfeld residual test. Pooled: fixed-effect inverse-variance estimate for FPC2; Het. $p$: Cochran's $Q$ test. Bold: $p < 0.05$.

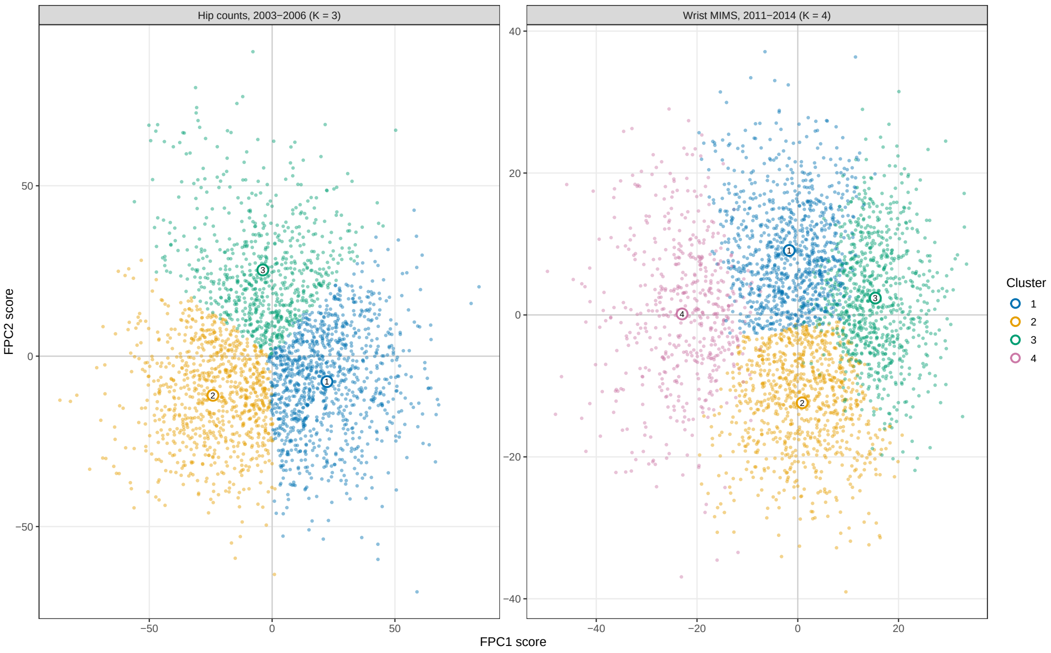


Figure S8: FPC1 and FPC2 scores coloured by the selected $k$-means solution, with cluster centres.

#### S2.7 Mortality prediction

Figure S10 shows forward selection of predictors of cardiovascular death when BMI, smoking, alcohol use, education and mobility difficulty were also candidates. Mobility difficulty was selected immediately after age by AIC and EPIC in both cohorts and by cross-validated AUC in the hip cohort (third in the wrist cohort). FPC2 was selected by cross-validated AUC and AIC in both cohorts and by EPIC in the wrist cohort, but not by EPIC in the hip cohort.

#### S2.8 Comparison with standard FPCA

Figure S11 extends the main-text comparison for FPC2 (Section 3.1.3) to all three residual components. Each residual component was re-estimated in 30 random samples of 300, 500, 1,000 and 2,000 participants per cohort and compared with its full-cohort version; the standard component at the rank matched in the full cohort (FPC5, FPC4 and FPC6) was compared with the same full-cohort residual component. Figure S12 shows one sample of 300 participants per cohort, chosen at the lower quartile of the similarity of standard FPC4 (a typical, not the worst, case).

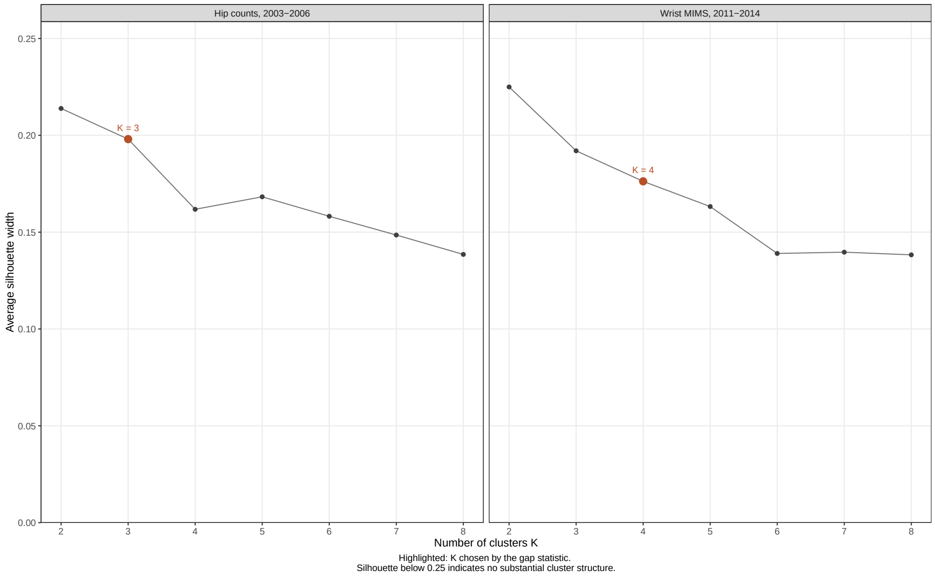


(a)

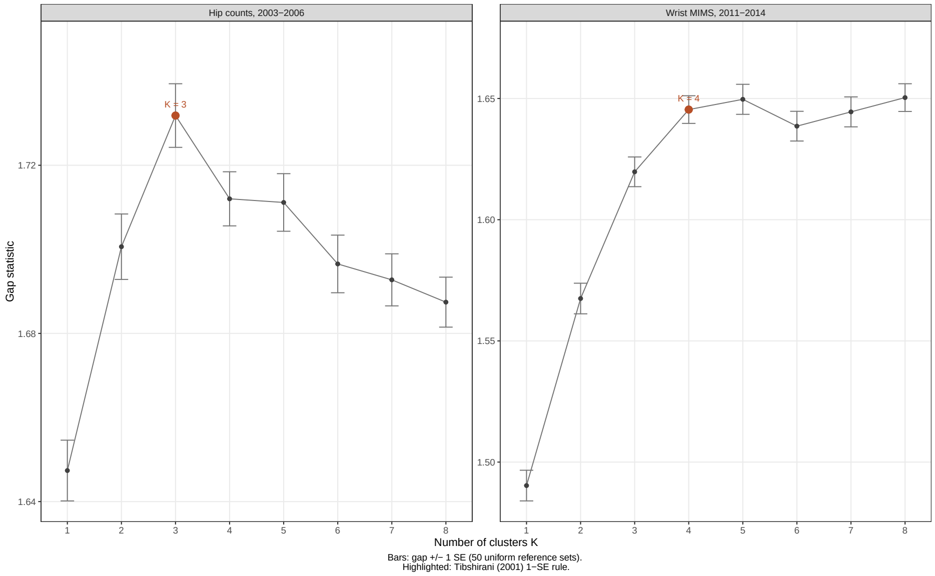


(b)

Figure S9: Clustering of the fPCA scores of the participant-average activity profiles. (a) Average silhouette width for $k$-means with $K = 2, \ldots, 8$; the highlighted point marks $K$ chosen by the gap statistic. (b) Gap statistic ($\pm 1$ SE) for $K = 1, \ldots, 8$; the highlighted point marks the $K$ selected by the 1-SE rule.

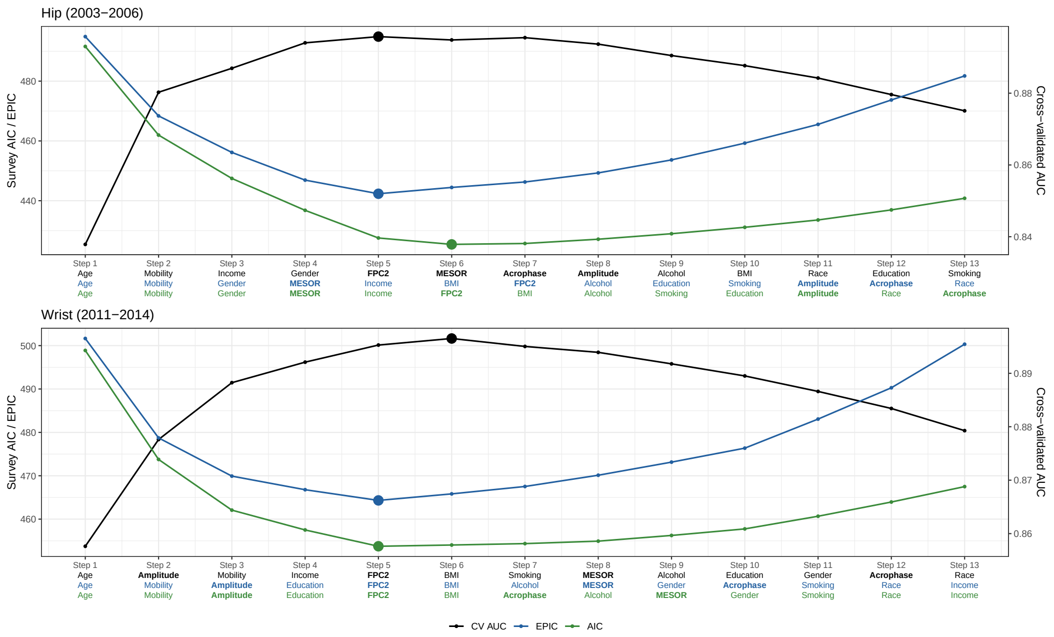

Figure S10: Forward selection of predictors of cardiovascular death with lifestyle and functional variables as additional candidates.

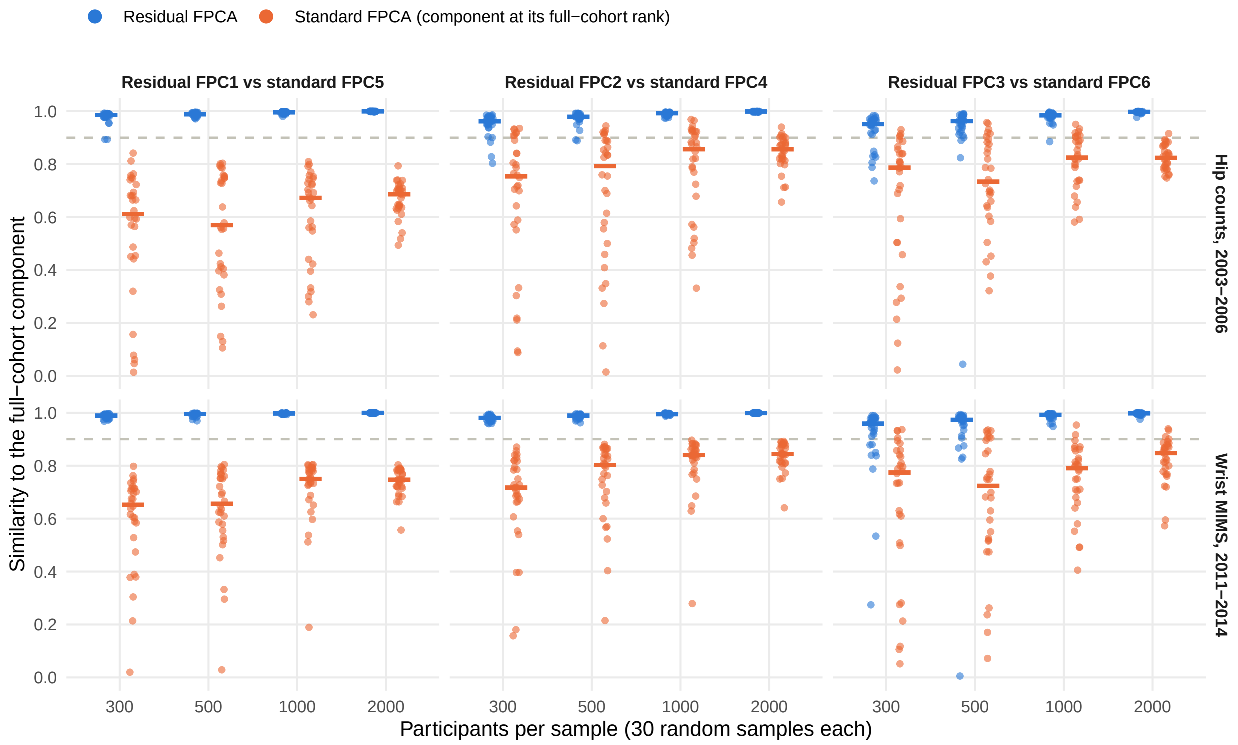

Figure S11: Stability of the residual components FPC1 to FPC3 (blue) and the matched standard component (orange). Points: individual samples; bars: median; dashed line: similarity 0.9.

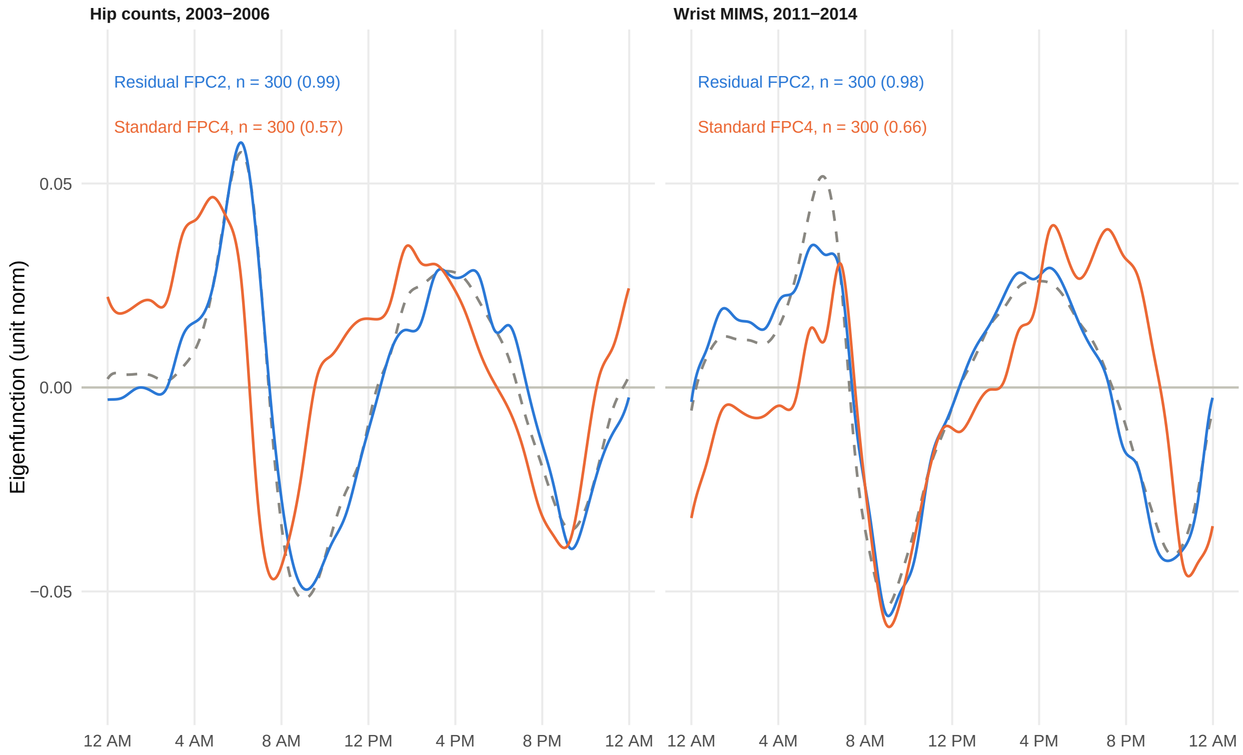


Figure S12: Residual FPC2 (blue) and standard FPC4 (orange) estimated in one random sample of 300 participants per cohort, against the full-cohort residual FPC2 (grey, dashed). Values in parentheses: similarity to the full-cohort residual FPC2. Signs were aligned with the full-cohort component.
