## Supplementary material for "Beyond the 24-Hour Rhythm: Functional Principal Components of Daily Activity Residuals and Mortality in NHANES": supp2

Xin Zheng*[1,2]

[1]Amsterdam UMC location Vrije Universiteit Amsterdam, Public and Occupational Health, De Boelelaan 1117, Amsterdam, the Netherlands
[2]Amsterdam Public Health, Health Behaviors and Chronic Diseases and Methodology, Amsterdam, the Netherlands

This section derives the properties of the cosinor-residual decomposition used in the main text. The results are standard properties of least squares and of covariance operators under linear maps; we state them for completeness and to make the conditions under which they hold explicit. All results below assume that every participant has complete, equally sampled days on the common grid.

#### 1 Notation

Each day is observed on a common grid of $n$ equally spaced times $t_1, \dots, t_n$ covering 24 hours (here $n = 1440$ minutes). Curves are written as vectors in $\mathbb{R}^n$, and the inner product $\langle f, g \rangle = \sum_{m=1}^{n} f(t_m) g(t_m)$ is the discrete analogue of $\int_0^{24} f(t)g(t)\,dt$. Let

$$X = \big[\,\mathbf{1},\ \cos(\omega t),\ \sin(\omega t)\,\big] \in \mathbb{R}^{n\times 3}, \qquad \omega = \frac{2\pi}{24\text{ h}}, \tag{1}$$

let $\mathcal{H}$ be the column space of $X$, and define

$$P_{\mathcal{H}} = X(X^\top X)^{-1}X^\top, \qquad Q = I - P_{\mathcal{H}}. \tag{2}$$

$P_{\mathcal{H}}$ is the orthogonal projection onto $\mathcal{H}$ and $Q$ the orthogonal projection onto its complement $\mathcal{H}^{\perp}$. Both are symmetric ($P_{\mathcal{H}}^{\top} = P_{\mathcal{H}}$, $Q^{\top} = Q$) and idempotent ($P_{\mathcal{H}}^2 = P_{\mathcal{H}}$, $Q^2 = Q$).

$X$ contains no participant data: it depends only on the time grid and the fixed 24-hour period. With complete data on a common grid, $X$, $P_{\mathcal{H}}$ and $Q$ are therefore identical for all participants. Participant-specific quantities are the daily curves $y_{ij}$, their average $\bar{y}_i = \frac{1}{7}\sum_j y_{ij}$, and the cosinor coefficients $\hat{\theta}_i = (M_i, \beta_i, \gamma_i)^{\top}$.

#### 2 The weekly fit equals the fit to the average day

**Proposition 1.** *With seven complete days on the common grid, the least-squares cosinor fit to all seven days equals the cosinor fit to the average day:* $\hat{\theta}_i = (X^{\top}X)^{-1}X^{\top}\bar{y}_i$.

*Proof.* The weekly fit minimises $\sum_{j=1}^{7}\|y_{ij} - X\theta\|^2$. Setting the gradient to zero gives the normal equations

$$\sum_{j=1}^{7} X^{\top}(y_{ij} - X\theta) = 0 \iff X^{\top}\Big(\sum_j y_{ij} - 7X\theta\Big) = 0 \iff X^{\top}(\bar{y}_i - X\theta) = 0, \qquad (3)$$

which are the normal equations for fitting $X\theta$ to $\bar{y}_i$. The days enter only through their average. □

The fitted curve is therefore $X\hat{\theta}_i = X(X^{\top}X)^{-1}X^{\top}\bar{y}_i = P_{\mathcal{H}}\,\bar{y}_i$, the projection of the average day onto $\mathcal{H}$.

#### 3 The average residual is the projection onto $\mathcal{H}^{\perp}$

**Proposition 2.** *The participant's average residual satisfies* $\bar{r}_i = Q\,\bar{y}_i$ *and* $X^{\top}\bar{r}_i = 0$.

*Proof.* Each daily residual is $r_{ij} = y_{ij} - X\hat{\theta}_i$. The fitted curve is the same on every day, so $\bar{r}_i = \frac{1}{7}\sum_j(y_{ij} - X\hat{\theta}_i) = \bar{y}_i - P_{\mathcal{H}}\bar{y}_i = Q\,\bar{y}_i$. Since $X^{\top}Q = X^{\top} - X^{\top}X(X^{\top}X)^{-1}X^{\top} = 0$, it follows that $X^{\top}\bar{r}_i = X^{\top}Q\,\bar{y}_i = 0$. □

Written out, $X^{\top}\bar{r}_i = 0$ is three conditions,

$$\sum_m \bar{r}_i(t_m) = 0, \qquad \sum_m \bar{r}_i(t_m)\cos(\omega t_m) = 0, \qquad \sum_m \bar{r}_i(t_m)\sin(\omega t_m) = 0, \qquad (4)$$

the discrete versions of the integral constraints: the average residual has zero mean and no 24-hour component, and so consists of the 12-hour and higher harmonics of the average day.

Intuitively, if the residual still contained a component along 1, $\cos(\omega t)$ or $\sin(\omega t)$, adjusting $M_i$, $\beta_i$ or $\gamma_i$ would reduce the residual sum of squares, contradicting optimality of the fit.

### 4 Covariance of the residuals

**Proposition 3.** *Let $\mathcal{C}_{\bar{y}}$ and $\mathcal{C}_{\bar{r}}$ be the covariance operators of $\bar{y}_i$ and $\bar{r}_i$ across participants. Then $\mathcal{C}_{\bar{r}} = Q\,\mathcal{C}_{\bar{y}}\,Q$.*

*Proof.* Because $Q$ is common to all participants, $\mu_{\bar{r}} = Q\mu_{\bar{y}}$ and $\bar{r}_i - \mu_{\bar{r}} = Q(\bar{y}_i - \mu_{\bar{y}})$. Hence

$$\mathcal{C}_{\bar{r}} = \mathbb{E}\big[(\bar{r}_i - \mu_{\bar{r}})(\bar{r}_i - \mu_{\bar{r}})^\top\big] = \mathbb{E}\big[Q(\bar{y}_i - \mu_{\bar{y}})(\bar{y}_i - \mu_{\bar{y}})^\top Q^\top\big] = Q\,\mathcal{C}_{\bar{y}}\,Q, \tag{5}$$

using that $Q$ is fixed and symmetric. This is the functional analogue of $\mathrm{Cov}(AY) = A\,\mathrm{Cov}(Y)A^\top$. □

### 5 Residual fPCA

#### 5.1 Residual eigenfunctions lie in $\mathcal{H}^\perp$

**Proposition 4.** *Every eigenfunction $\varphi$ of $\mathcal{C}_{\bar{r}}$ with eigenvalue $\lambda > 0$ lies in $\mathcal{H}^\perp$; in particular $\sum_m \varphi(t_m) = \sum_m \varphi(t_m)\cos(\omega t_m) = \sum_m \varphi(t_m)\sin(\omega t_m) = 0$.*

*Proof.* From $\mathcal{C}_{\bar{r}}\varphi = \lambda\varphi$ with $\lambda > 0$, $\varphi = \lambda^{-1}Q\,\mathcal{C}_{\bar{y}}\,Q\,\varphi$, which is in the range of $Q$, i.e. in $\mathcal{H}^\perp$. Hence $\varphi$ is orthogonal to each of 1, $\cos(\omega t)$ and $\sin(\omega t)$. Orthogonality to the constant function, $\cos(\omega t)$ and $\sin(\omega t)$ is the zero-sum (zero-integral) condition. □

#### 5.2 Relation to fPCA of the activity curves

Eigenfunctions of $\mathcal{C}_{\bar{y}}$ are mutually orthogonal, as for any symmetric operator, but nothing constrains them to be orthogonal to $\mathcal{H}$. Writing $\bar{y}_i = P_{\mathcal{H}}\bar{y}_i + Q\bar{y}_i$ gives

$$\mathcal{C}_{\bar{y}} = \underbrace{P_{\mathcal{H}}\mathcal{C}_{\bar{y}}P_{\mathcal{H}}}_{\text{cosinor part}} + \underbrace{Q\,\mathcal{C}_{\bar{y}}\,Q}_{=\mathcal{C}_{\bar{r}}} + \underbrace{P_{\mathcal{H}}\mathcal{C}_{\bar{y}}Q + Q\,\mathcal{C}_{\bar{y}}P_{\mathcal{H}}}_{\text{cross terms}}. \tag{6}$$

The cross terms are the covariance, across participants, between the cosinor part and the residual part of the average day.

- If the cross terms vanish, $\mathcal{C}_{\bar{y}}$ is block diagonal: each of its eigenfunctions lies entirely in $\mathcal{H}$ or entirely in $\mathcal{H}^\perp$, and those in $\mathcal{H}^\perp$ are the eigenfunctions of $\mathcal{C}_{\bar{r}}$. Raw and residual fPCA then agree, up to the ordering of components.

- If the cross terms are non-zero, eigenfunctions of $\mathcal{C}_{\bar{y}}$ combine 24-hour and higher-harmonic variation, and in general none coincides with an eigenfunction of $\mathcal{C}_{\bar{r}}$. Discarding the leading components of fPCA on $\bar{y}_i$ is then not equivalent to fPCA of $\bar{r}_i$.

#### 5.3 Orthogonal functions, not necessarily uncorrelated scores

The constraints above apply to each curve over time, not across participants: shape scores may still be correlated with mesor, amplitude or acrophase, as reflected by the cross terms in (6). Association analyses therefore adjust for these parameters.

#### 5.4 Day-level residuals

For the daily residuals, $r_{ij} - \bar{r}_i = (y_{ij} - X\hat{\theta}_i) - (\bar{y}_i - X\hat{\theta}_i) = y_{ij} - \bar{y}_i$. Within-participant deviations are therefore not projected off $\mathcal{H}$: with a weekly cosinor fit, the level-2 components of the multilevel fPCA can include day-to-day variation in level, amplitude and timing as well as shape. Level-1 (between-participant) components describe shape beyond each participant's own cosinor rhythm.
